# Potential clinical impact of predictive modeling of heterogeneous treatment effects: scoping review of the impact of the PATH Statement

**DOI:** 10.1101/2024.05.06.24306774

**Authors:** Joe V Selby, Carolien C H M Maas, Bruce H Fireman, David M Kent

## Abstract

**Background:** The PATH Statement (2020) proposed predictive modeling for examining heterogeneity in treatment effects (HTE) in randomized clinical trials (RCTs). It distinguished risk modeling, which develops a multivariable model predicting individual baseline risk of study outcomes and examines treatment effects across risk strata, from effect modeling, which directly estimates individual treatment effects from models that include treatment, multiple patient characteristics and interactions of treatment with selected characteristics.

**Purpose:** To identify, describe and evaluate findings from reports that cite the Statement and present predictive modeling of HTE in RCTs.

**Data Extraction:** We identified reports using PubMed, Google Scholar, Web of Science, SCOPUS through July 5, 2024. Using double review with adjudication, we assessed consistency with Statement recommendations, credibility of HTE findings (applying criteria adapted from the Instrument to assess Credibility of Effect Modification Analyses (ICEMAN)), and clinical importance of credible findings.

**Results:** We identified 65 reports (presenting 31 risk models, 41 effect models). Contrary to Statement recommendations, only 25 of 48 studies with positive overall findings included a risk model; most effect models included multiple predictors with little prior evidence for HTE. Claims of HTE were noted in 23 risk modeling and 31 effect modeling reports, but risk modeling met credibility criteria more frequently (87 vs 32 percent). For effect models, external validation of HTE findings was critical in establishing credibility. Credible HTE from either approach was usually judged clinically important (24 of 30). In 19 reports from trials suggesting overall treatment benefits, modeling identified subgroups of 5-67% of patients predicted to experience no benefit or net treatment harm. In five that found no overall benefit, subgroups of 25-60% of patients were nevertheless predicted to benefit.

**Conclusions:** Multivariable predictive modeling identified credible, clinically important HTE in one third of 65 reports. Risk modeling found credible HTE more frequently; effect modeling analyses were usually exploratory, but external validation served to increase credibility.

## INTRODUCTION

Overall, or average, treatment effects from randomized clinical trials (RCTs) provide limited information to patients making personal treatment decisions. (1–6) Even in strongly positive RCTs, some patients do not benefit from the favored treatment and yet may experience adverse effects. Patients and clinicians would benefit greatly if more individualized evidence could be generated and reported from RCTs.

Most publications of RCT results continue to limit examination for possible heterogeneity of treatment effects (HTE) to one-at-a-time comparisons between numerous patient subgroups, e.g. men vs. women, persons with vs. without diabetes, even though guidelines for identifying HTE in RCTs (1,7–11) have long emphasized that such analyses are at great risk for both false positive and false negative findings. An additional limitation of this approach is that individuals simultaneously belong to multiple subgroups that may vary in whether or how they appear to benefit. Thus, guidelines consistently recommend limiting the number of subgroups studied to those with prior evidence or strong biologic or clinical rationale for HTE and using caution in interpreting or applying findings to clinical practice.

The emergence of precision medicine (12) and patient-centered outcomes research (13) heightens interest in identifying important HTE. In 2020, an expert panel funded by the Patient-Centered Outcomes Research Institute (PCORI) published The Predictive Approaches to Treatment Heterogeneity (PATH) Statement (14,15), which described predictive modeling approaches that incorporate multiple patient attributes simultaneously to identify HTE and predict individualized treatment effects. The Statement pointed out that HTE, sometimes referred to as treatment effect modification and tested as a statistical interaction, should be sought on both the absolute scale (e.g., as risk *differences)* and on the relative scale (e.g., as *ratios* of risks, odds or hazards) and emphasized that heterogeneity in absolute treatment effects matters more to individual patients and clinicians making treatment decisions.

The Statement distinguished two approaches to predictive modeling. “Risk modeling” incorporates multiple baseline patient characteristics into a model predicting risk for the RCT’s outcome (usually the primary outcome). In a second step, both absolute and relative treatment effects are examined across pre-specified strata (e.g., quarters) of predicted risk. (16) In the second approach, “effect modeling”, a model is developed within the RCT data to directly estimate individual treatment effects by including treatment, multiple covariates and interactions of treatment with one or more covariates. Both regression methods and more flexible, non-parametric, data-driven machine-learning algorithms (e.g., 17-20) have been used in effect modeling.

The Statement recommended risk modeling whenever an RCT demonstrates an overall treatment effect. Risk of study outcomes varies substantially in most RCT populations when participants are stratified using a multivariable model. Assuming that the relative treatment effect is homogeneous in the population, the absolute benefit would be expected to increase as predicted risk increases, (21–22) This mathematical relationship has been called “risk magnification.” (23) and is often referenced in the evidence-based medicine literature (10,24,25) and is implicit in clinical guidelines that reserve treatments with high costs or potential adverse effects for those with higher baseline risk (26,27) The Statement encouraged use of validated external models for predicting risk if available, but indicated that in their absence, models can be developed within the RCT population, using baseline covariates and observed study outcomes and including both study arms.

Although effect modeling theoretically permits a more robust examination of possible HTE, the Statement emphasized the vulnerability of these approaches to overfitting, (28) and recommended that effect modeling be used only when there are a small number of previously established effect modifiers. It also encouraged use of methods to reduce risks of “over-fitting” of data as well as validation of effect model findings in external datasets when possible.

We conducted a scoping review (29) to assess the impact of the PATH Statement in terms of the frequency with which predictive modeling analyses of RCT data have appeared and cited the PATH Statement since its publication, the consistency of analyses with Statement consensus criteria (Supplement Table 2), and the credibility and clinical importance of claimed HTE. We applied criteria adapted from the Instrument to assess Credibility of Effect Modification Analyses (ICEMAN) (11) to assess credibility, and when HTE was found to be credible, we assessed “clinical importance,” using the Statement definition of “variation across patients in the absolute treatment effect sufficient to span clinically-defined decision thresholds, supporting differing treatment recommendations for patient subgroups.”

## METHODS

### Identification of Reports for Inclusion

Using the Cited By functions in PubMed, Google Scholar, Web of Science and the SCOPUS database (Supplement Table 1A), we identified reports that appeared between January 7, 2020 and July 24, 2024, cited the Statement and presented multivariable predictive modeling in RCT data to identify HTE. We included non-peer-reviewed reports from pre-print archives and dissertations on institutional websites.

Among 312 citations identified (Figure 1), 83 (30–112) involved analyses of data from RCTs. Sixteen (30–45) were excluded because they did not examine HTE using multivariable predictive models. Supplement Table 1B presents reasons for exclusion. Three analyses were represented by two reports each (46–51) and one report (110) presented predictive models from two distinct trials in different clinical areas, leaving 65 reports analyzing data from 162 RCTs.

**Figure 1.**
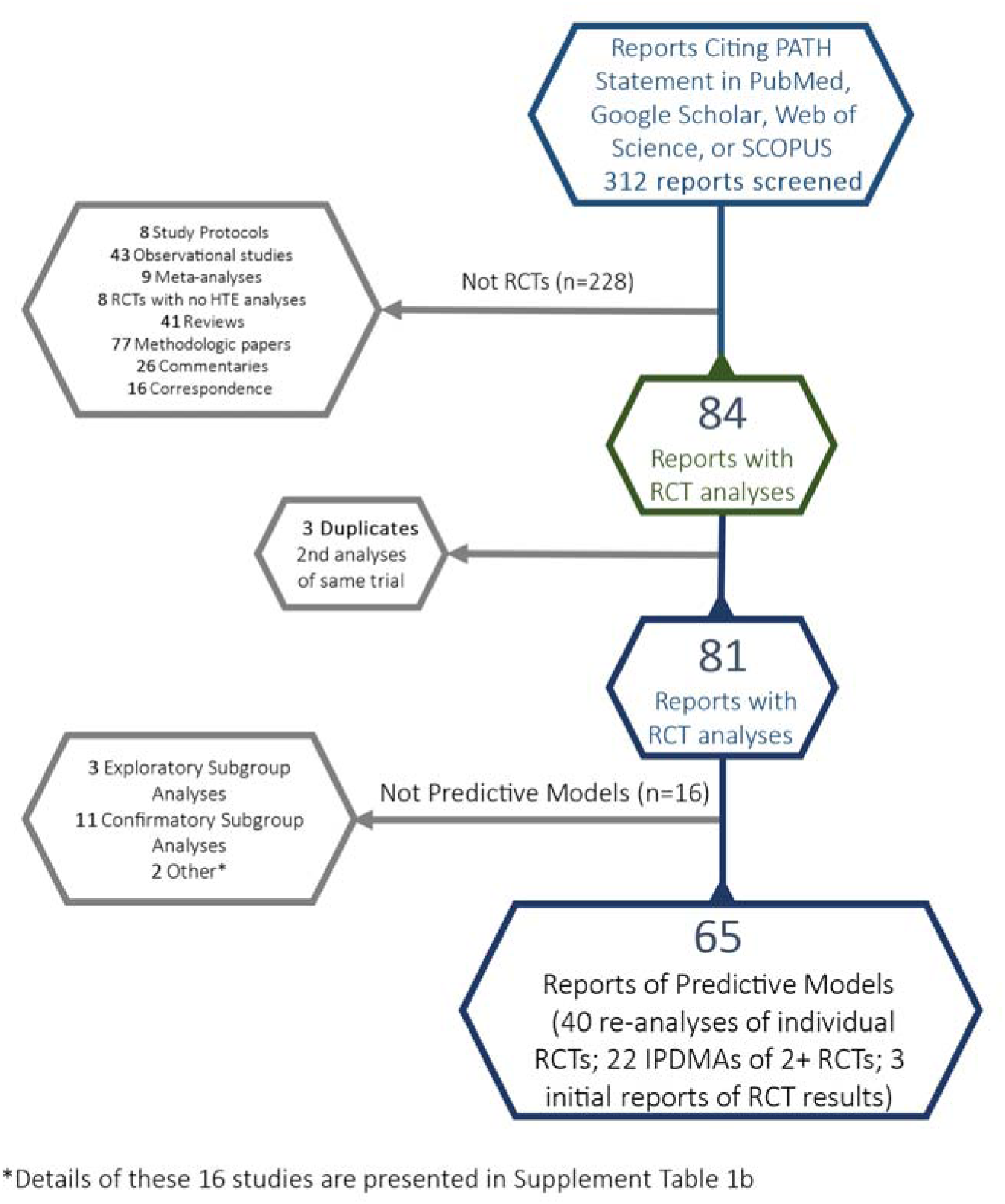
Flow Diagram for identification and screening of all reports citing the PATH Statement and for exclusion of reports not meeting study criteria for presenting a predictive model of individual treatment effects from RCT data. Abbreviations: RCT: randomized controlled trial; IPDMA: independent patient data meta-analysis.

### Review of Predictive Model Reports

Variables collected and coding instructions are presented in Supplement Tables 3 and 4, respectively. All details of analytic strategies and findings of HTE were doubly reviewed by the first author and one co-author. A “learning” set of six reports was reviewed and discussed by all co-authors. Thereafter, co-reviewers discussed and resolved initial disagreements.

Review classified each report as risk modeling, effect modeling or both and further classified effect models into those based primarily on regression methods (e.g., ordinary least squares, logistic, proportional hazards, Bayesian regression methods) and those using more flexible data-driven machine-learning algorithms. For both risk and effect modeling analyses, we noted whether authors reported having found HTE on either absolute or relative scales and whether results of statistical testing for HTE were presented. We included statistical tests for treatment-covariate interactions from regression models, direct contrasts of treatment effects across subgroups, reporting of confidence intervals for subgroup treatment effect estimates, and overall tests of the null hypothesis of homogeneity of treatment effect in machine-learning algorithms.

We determined whether the performance of final models for predicting individual or subgroup treatment effects was validated in datasets external to the derivation population, including validations conducted in entirely distinct RCTs, those conducted in pre-specified, non-random subsets of the original RCT population (e.g., subsets selected on bases of geography (trial sites) or time of enrollment), and those conducted in large observational cohorts.

#### Assessment of Credible and Clinically Important HTE

To assess credibility of claimed HTE, on either absolute or relative scales, we adapted four of the five ICEMAN criteria for RCTs (11,113). Detailed description and scoring guidance for each criterion and the overall credibility score are detailed in Supplement Tables 7A and 7B. Although ICEMAN criteria were originally developed for evaluating treatment effect modification by single covariates, four apply readily to predictive modeling with multiple covariates. These include 1) Did the authors test only a small number of interactions; 2) Was possible effect modification by each covariate supported by prior evidence; 3) If the covariate is a continuous variable, were arbitrary, data-driven cut points avoided; and 4) Does a statistical test for interaction suggest that chance is an unlikely explanation of the apparent HTE? The fifth criterion, whether the direction of interaction was hypothesized in advance, is not applicable to predictive modeling, given that multiple covariates and potentially complex interactions are evaluated simultaneously. No single criterion, including that of statistical testing, is treated as either sufficient or necessary for establishing overall credibility. Overall credibility scores range from 1 to 4 (very low, low, moderate, or high credibility).

Because risk models involve a single effect modifier (the baseline risk score) with strong prior theoretical and empirical support (21) for HTE, at least on the absolute scale; and because risk modeling either reports pre-specified risk score cut-points or treats risk as a continuous variable, risk models can be expected to score well when ICEMAN criteria are applied. Most effect models tested multiple potential treatment-covariate interactions, often with little prior evidence and therefore tended to score poorly with application of adapted ICEMAN criteria. However, we gave considerable weight to external validation of effect model performance in another population. External validation essentially tests for HTE across a single vector, the “effect score” or predicted individual treatment effect, much as risk modeling tests for HTE across the risk score. (114) Overall credibility usually rose to “moderate” if models performed well in external validation, even if credibility of derivation analyses would have been scored as very low.

We classified all reports scored as at least moderate overall credibility as “credible” and assessed findings for clinical importance. Per the PATH Statement, clinical importance is based exclusively on the size and direction of observed differences in absolute treatment effects between subgroups and whether these differences appear sufficient to support differing treatment recommendations. An additional consideration was whether findings for all outcomes studied, including adverse effects of treatment, were consistent in supporting the same treatment choice.

## Results

### General Description

Predictive models of HTE appeared with increasing frequency each year following publication of the PATH Statement (Table 1). Among the 65 reports (46–112), we identified 31 risk modeling and 41 effect modeling analyses. Seven reports (60,84,86,91,105,109,111) presented both risk and effect modeling analyses. Most effect modeling reports examined large numbers of potential interactions, and the majority employed data-driven, non-parametric analytic methods.

**Table 1.**
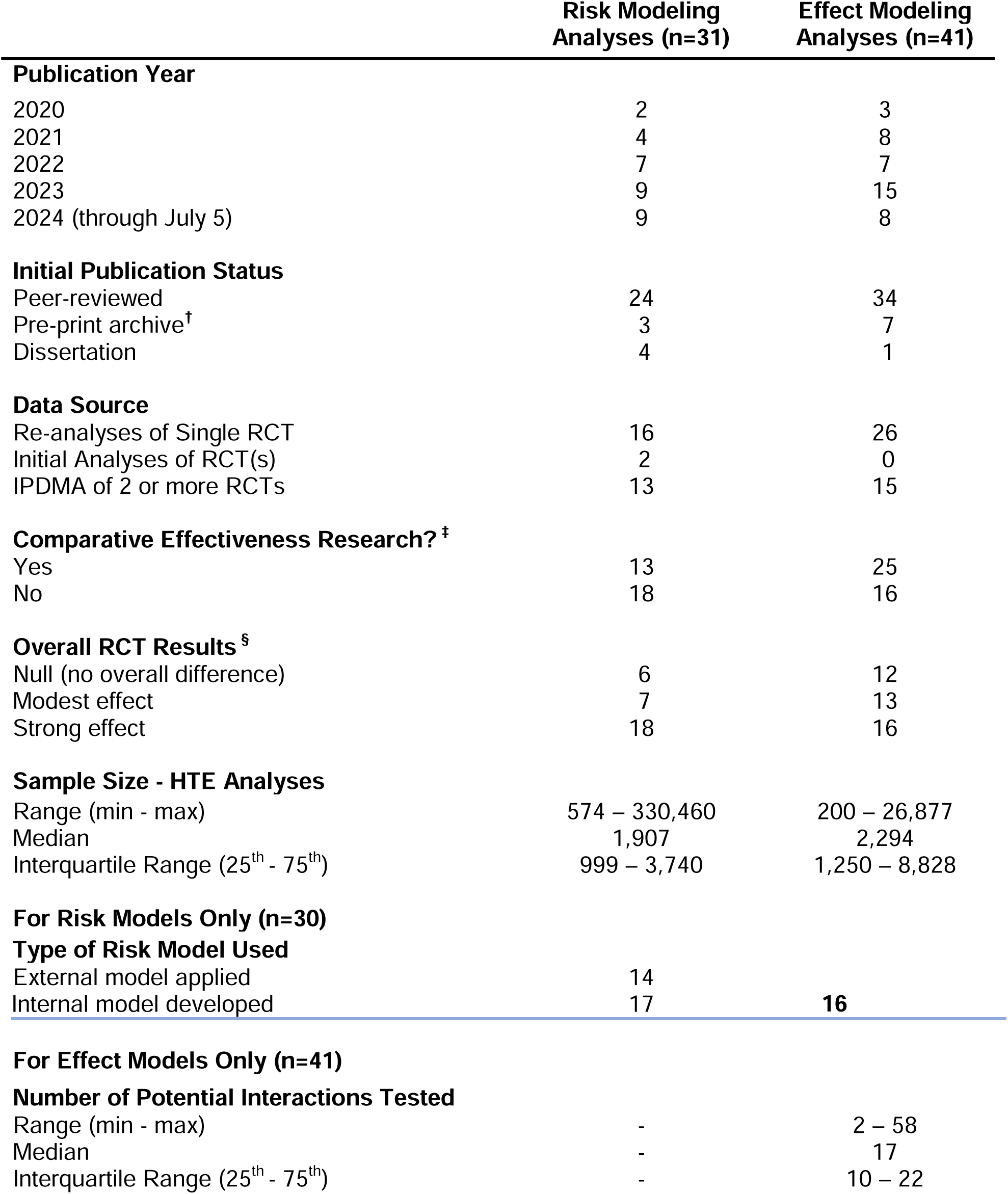
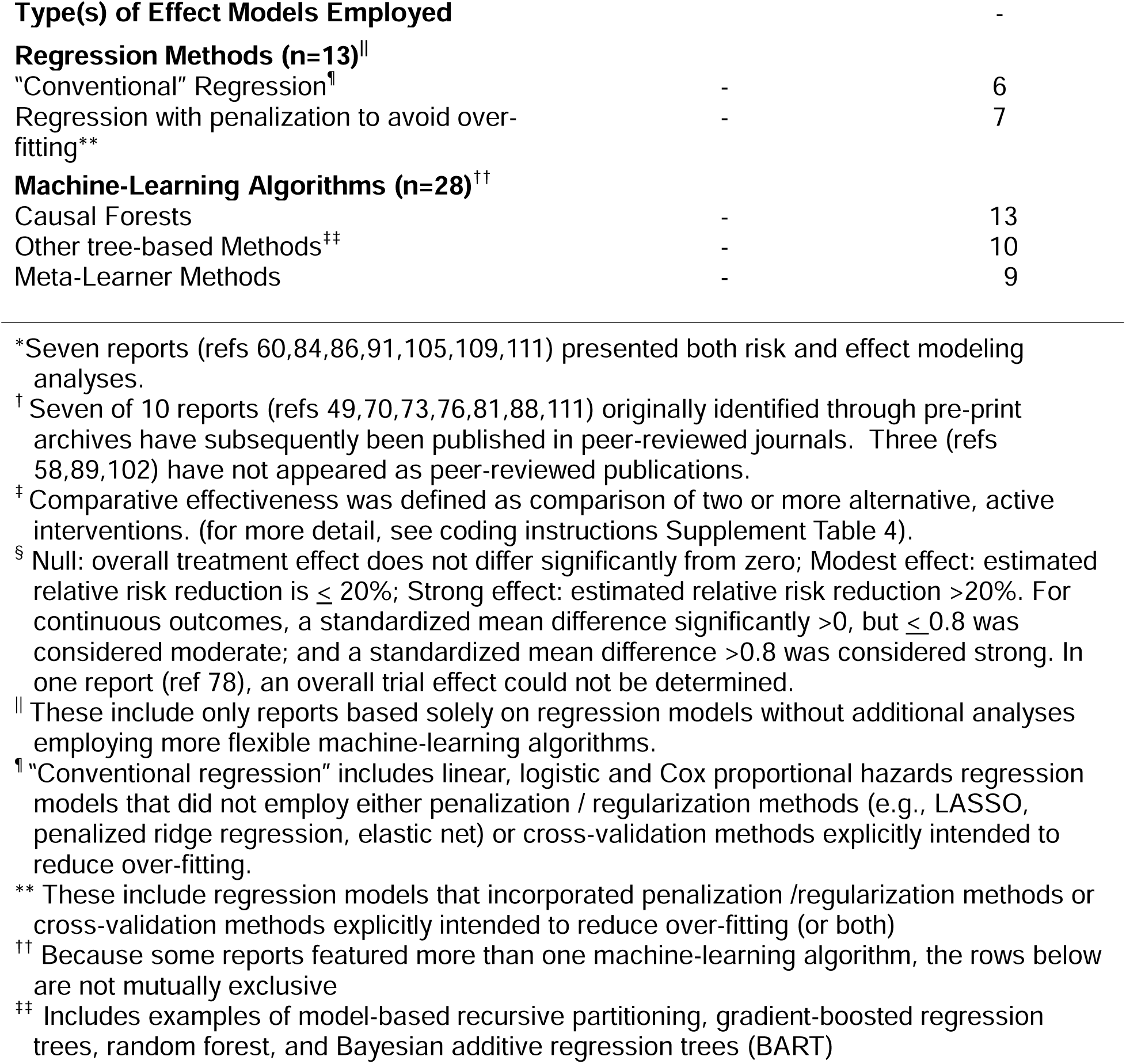
Characteristics of the 70 analyses (65* reports) of predictive models for possible heterogeneity of treatment effect.

### Reviewer Agreement

Excluding six reports (presenting six effect models and one rik model) used for training reviewers, initial between-reviewer disagreement rates for 19 doubly-reviewed items ranged from 0 to 47%, with an overall average of 10.1% (details, Supplement Table 5). Initial disagreement was greater for assessments of the credibility and clinical importance of claimed HTE. Possibly because assessment of credibility and clinical importance were added near the end of data collection and without additional training, initial disagreement was more common for these items, although generally this was easily resolved upon discussion.

### Risk Models

Concordance with eight Statement criteria related to risk modeling was above 60% for 5 of 8 (Supplement Table 3). Only 52% (25) of the 48 reports with positive overall findings included a risk model. Fourteen of 31 risk modeling analyses used an external prediction model. Half of reports presented risk model scores by treatment arm. All but one presented absolute treatment effects by level of risk and most reported relative treatment effects as well.

Study authors claimed findings of HTE in 23 of 31 risk modeling analyses (Figure 2, Supplement Table 8). For 13 reports, HTE was found on the absolute but not the relative scale (i.e., risk magnification). For the remaining 10, relative treatment effects also appeared to vary across levels of baseline risk. In five, (49,61,79,91,96) relative treatment effects were greater in individuals at higher risk for experiencing trial outcomes. Relative treatment benefit was confined to individuals in the middle of the risk distribution in three reports (47,65,69) or to those at lowest risk in two (59,84).

**Figure 2.**
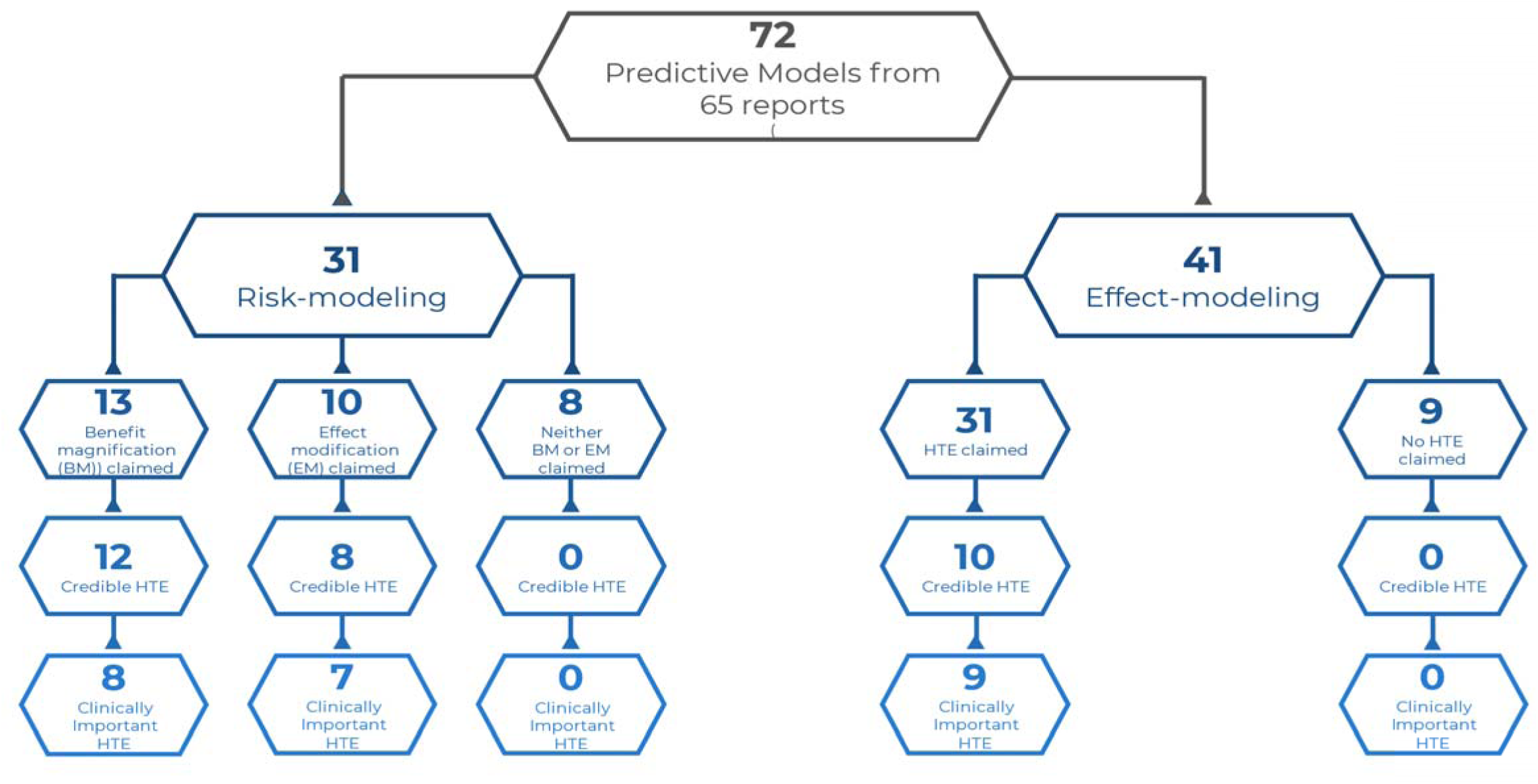
Adjudicated results of review of all eligible reports for type of predictive modeling (risk or effect), for claims by authors of heterogeneity of treatment effects (HTE), for credibility of HTE (using adapted ICEMAN criteria), and for clinical importance of HTE found to be credible.

### Effect Models

Concordance was low for 3 of the 6 Statement criteria among effect modeling reports (Supplement Table 4). Only 6 of 41 (52,60,84,86,108,109) restricted analyses to small numbers of covariates with strong prior evidence for effect modification. Most explored many candidate effect modifiers with little prior evidence. Only nine (51,52,53,55,70,80,92,93,108) applied effect model findings to external datasets for validation. Authors claimed HTE in 31 of the 41 effect model reports (Figure 2, Supplement Table 8). Thirty presented evidence for absolute treatment effect differences across subgroups; nine presented evidence for relative effect differences. In 14 of the 31, authors heeded recommendations to report model performance metrics that evaluate prediction of individual treatment effects rather than prediction of risk for the outcome.

### Assessment for Credibility of HTE

Most reports, whether of risk or effect modeling, claimed to have identified HTE (Figure 2), but as expected, risk models were more likely to be scored as credible when ICEMAN criteria were applied. Detailed scores for the 51 reports claiming HTE are given in Supplement Table 8. Twelve of 13 risk modeling analyses claiming risk magnification and eight of ten that found HTE on a relative scale were scored as credible. By contrast, findings of HTE in 31 effect modeling reports were judged credible in only 10. Most effect modeling reports explored many variables with little prior evidence for effect modification. Those employing data-driven machine-learning algorithms also allowed the data to guide cut-point selection for continuous covariates. Among the 10 effect modeling reports judged to present credible HTE, nine (51,52,55,70,80,92,93,108,111) validated model predictions of individual treatment effects in independent cohorts, usually another RCT. Three of the ten (52,99,108) also met ICEMAN criteria (and PATH recommendations) by restricting analyses to a very small number of candidate effect modifiers with strong prior evidence.

### Assessment for Clinically Important HTE

Reviewers judged findings from 24 of 30 reports with credible HTE to be clinically important (Figure 2). Details for these are 24, including rationales for classification as clinically important, are summarized in Table 2. In 19, RCT findings had suggested a moderate (n=6) or strong (n=13) benefit of one treatment vs. another. Yet, predictive modeling identified subgroups representing 5 to 67% of the trial population for whom no benefit or possible net harm would be expected from that treatment. In five, overall results suggested no benefit, but predictive modeling identified subgroups representing 25-67% of participants who did appear to benefit from one treatment vs. another. Findings of six reports with credible HTE were judged not clinically important because the heterogeneity, though credible, did not span a threshold suggesting differing treatment choices; (95,112) or because of conflicting findings across outcomes, (75,88) failure to add clinical value to previous risk-based selection strategies, (79) or concurrence with authors on the need for additional investigation, possibly testing additional effect modifiers. (55)

**Table 2.** Studies found to have credible and clinically important heterogeneity of treatment effects (HTE)

| Reference | Clinical Condition | Outcomes | Randomized Intervention (s) | Overall RCT Findings | Methods | Type of HTE Identified | Clinically Important Subgroup Differences |
| --- | --- | --- | --- | --- | --- | --- | --- |
| **RISK MODELS (n=15)** |  |  |  |  |  |  |  |
| Redelmeier et al.<sup>46,47</sup> | Congestive heart failure | All-cause mortality | Implantable defibrillator (ICD) vs medical management | Strong benefit in favor of ICD (OR=0.69) | Single RCT re-analysis; internal risk model | Relative as well as absolute effect heterogeneity, $p < 0.001$ for multiplicative interaction | Despite a strong overall benefit of ICD implantation, the mortality benefit was largely confined to patients in midrange of risk (3<sup>rd</sup> and 4<sup>th</sup> quintiles), i.e., a “sweet spot.” Intervention associated with high costs, patient discomfort. |
| Chalkou et al.<sup>48,49</sup> | Multiple sclerosis (MS) | Relapse of MS | 3 immunologic therapies (DF: Dimethyl fumarate, GA: Glatiramer acetate, N: Natalizumab) vs placebo | Strong benefit in favor of Natalizumab (OR<sub>vs placebo</sub>: DF: 0.43<br>GA: 0.53<br>N: 0.28) | IPDMA – 3 RCTs, using network meta-analysis; external risk model | Probable relative as well as absolute effect heterogeneity; test for multiplicative interaction not reported | Despite findings of strong overall benefit of natalizumab, patients with baseline risk < 30%, comprising 25% of trial population, had negligible added benefit of natalizumab vs. dimethyl fumarate. Natalizumab is associated with rare but possibly fatal complication, progressive multifocal leukoencephalopathy (PML). |
| Kumar et al.<sup>54</sup> | Ambulatory patients with cancer | Venous thrombo-embolism (VTE) | Apixaban vs placebo | Strong benefit in favor of apixaban (aHR=0.49) | Single RCT re-analysis; external risk model | Probable relative as well as absolute effect heterogeneity; test for multiplicative interaction not reported | Despite a strong overall benefit of apixaban, patients with a baseline risk for VTE < 8% (67% of the trial population) derived no benefit, but experienced an excess of overall bleeding events on treatment with apixaban |
| Bress et al.<sup>60</sup> | Systolic hypertension and increased cardiovascular risk | Cardiovascular disease events; all-cause mortality | Intensive vs standard systolic blood pressure control | Strong benefit in favor of intensive systolic control (HR=0.75 and 0.73, respectively) | Single RCT re-analysis; internal risk model | Absolute but not relative effect heterogeneity (risk magnification) | Despite the strong overall benefit of intensive SBP control, patients in lowest risk quartile would expect very little benefit from intensive control, despite increased costs, burden and adverse effects. |

| Reference | Clinical Condition | Outcome(s) | Randomized Interventions | Overall RCT Findings | Methods | Type of HTE Identified | Clinically Important Subgroup Differences |
| --- | --- | --- | --- | --- | --- | --- | --- |
| Rysavy et al.<sup>59</sup> | Extreme Prematurity | Broncho-pulmonary dysplasia | Vitamin A vs sham injection | Weak benefit in favor of vitamin A (RR=0.89) | Single RCT re-analysis; external risk model | Relative as well as absolute effect heterogeneity, p=0.03 for multiplicative interaction | In the face of a weak overall benefit of Vitamin A therapy, predictive modeling found a strong benefit in the 50% of infants at lower predicted risk; no suggestion of benefit among infants in highest quarter of predicted risk. |
| Kent et al.<sup>61</sup> | Patent foramen ovale (PFO) - associated stroke | Recurrent stroke | Percutaneous PFO-closure vs medical therapy | Strong benefit in favor of PFO-closure (aHR=0.41) | IPD meta-analysis of 6 RCTs; external risk model | Relative as well as absolute effect heterogeneity, p=0.003 for multiplicative interaction | Despite a strong overall benefit, analyses identified a subgroup comprising 15% of trial population unlikely to have had a PFO-related stroke. This subgroup received no benefit from PFO-closure and were at higher risk of procedure-related complications. |
| Taylor et al.<sup>69</sup> | Hospitalized Patients with Sepsis | 30-day mortality and readmission | Nurse-navigator led Sepsis Transition and Recovery (STAR) intervention vs usual care | Weak benefit in favor of the intervention (aOR=0.80) | Single RCT re-analysis; external risk model | Relative as well as absolute effect heterogeneity; 95% confidence intervals for quartile-specific OR's do not overlap | Despite findings of a modest overall benefit of the program, the benefit was confined to patients in the midrange of predicted risk (middle two quartiles) i.e., "sweet spot"). Intervention is associated with greater costs. |
| Gencer et al.<sup>71</sup> | Atrial Fibrillation | Net composite of stroke/ systemic embolism, major bleed, all-cause death | Lower (LDER) vs higher (HDER) dose regimens of edoxaban vs warfarin | Weak benefit for either dose of edoxaban vs warfarin (HR=0.83, 0.89 for LDER, HDER respectively) | Single RCT re-analysis; Risk stratification based on count of number of high-risk features | Absolute effect heterogeneity (p=0.001) but probably not relative effect heterogeneity (p=0.065) (risk magnification) | With a weak overall effect, the absolute benefits of either dose of edoxaban vs warfarin were shown to be negligible in those with 0-1 risk factors, but to increase for both stroke/embolism and major bleeding endpoints as number of risk factors increases. May not justify switching in low-risk patients well managed on warfarin. |
| Trinks-Roerdink et al.<sup>82</sup> | Atrial Fibrillation | All-cause mortality | Integrated atrial fibrillation care vs. usual care | Strong benefit of integrated care (aHR=0.55) | Single RCT re-analysis, external risk model | Absolute but no relative effect heterogeneity (p=0.93 for multiplicative interaction) (risk magnification) | Despite a strong overall benefit of the program, benefits were shown to be predominantly in persons in highest 25% of risk distribution; minimal benefit with increased increased costs in lower 75%. |
| Goligher et al.<sup>84</sup> | Hospitalized COVID-19 infection | Organ-support free days; hospital survival | Therapeutic-dose heparin vs usual pharmacologic thromboprophylaxis | Null (No overall benefit in population (OR for benefit 1.05) | IPDMA – 3 trials; re-analysis; subgroup analyses, internal risk model and effect model) | Apparent qualitative interaction on both relative and absolute scales; statistical significance not tested for risk model | Although no overall benefit was observed, a clear benefit was shown for the 60% of persons in lower deciles of risk score (not requiring organ support at baseline); apparent harm for those needing intensive care at baseline (highest 3 deciles of risk score); note: similar HTE was suggested in subgroup analyses and in an effect model |
| Xu et al.<sup>94</sup> | Healthy people 70+ yrs old; | Composite of death, dementia or disability | Low-dose (100mg) aspirin vs placebo | Null (HR=1.01, 95%CI: 0.92-1.11) | Single RCT re-analysis, external risk model | Absolute effect heterogeneity (p=0.03), but no relative effect heterogeneity (risk magnification) | Despite a null overall effect, risk model showed that highest risk 20% of patients benefitted (ARR = 15.1%; 95% CI 4.0–26.3%), with little suggestion of any benefit in remaining 80%. |
| Paules et al.<sup>96</sup> | Patients hospitalized with Covid-19 | 28-day mortality, hospitalization, progression to mechanical ventilation or death; recovery rate. | Baricitinib + remdesivir vs. remdesivir alone | Strong Benefit of combination for each outcome. (HR for death, 0.65; 95% CI, 0.39 to 1.09) | Single RCT re-analysis, external risk model | Clear heterogeneity on absolute scale for all 3 outcomes (risk magnification); also, possible relative effect heterogeneity for the time-to-recovery outcome, with HR for for the highest risk quartile significantly greater than for lowest risk quartile | A strong overall benefit of baricitinib in the trial was shown to be due to a very strong benefit on absolute and relative scales in the highest risk quartile, and likely the 2<sup>nd</sup> highest risk quartile as well; with little apparent effect in the lower 50% of predicted risk. |
| Vickers et al.<sup>97</sup> | Localized prostate cancer | 15-year prostate cancer mortality | Radical prostatectomy vs conservative care | Strong benefit of surgery (HR=0.55) | Separate RCT re-analyses from two trials, external risk models | Established that HR's were constant across levels of baseline risk (p-values for interaction on relative scale >0.20 in both trials); therefore: risk magnification | Despite a strong overall relative benefit, the authors show that absolute benefit of radical prostatectomy in the lowest risk quartile of current cohorts would be approximately a 0.2% reduction in 15-year mortality. For higher risk subgroups, benefit is substantial. This is useful information for shared decision-making |
| De Winkel et al.<sup>109</sup> | Acute subarachnoid hemorrhage | Functional status at 2 mos; re-treatment or re-bleed by 10 yrs. | Endovascular coiling vs intracranial clip | Strong benefit of coiling for good functional status (OR for benefit: 1.5, 95% CI 1.3, 1.7); but a strong benefit of clipping for 10-yr treatment failure: (HR: 0.30, 95% CI 0.2, 0.4) | Single RCT re-analysis; ordinal regression for functional status; proportional hazards model for treatment failure; began as exploratory effect modeling but reduced to risk modeling | Established strong risk magnification favoring intracranial clip for outcome of re-bleed or re-bleed. Modest differences in absolute benefits of coiling across range of risk for poor functional outcome | Despite competing overall findings for the two outcomes, indicating need for shared decision-making, a subgroup of 6% were predicted to have a strong benefit of clipping for the re-bleed/retreatment outcome with very low risk and negligible (<2%) differences between treatments for the functional status outcome. |
| Smit et al.<sup>111</sup> | Community-acquired pneumonia | 30-day mortality | Corticosteroids vs placebo | Strong benefit of corticosteroids (OR 0.72; 95% CI 0.56-0.94) | IPDMA of 6 trials, external risk models; also examined an effect model (see below) | No evidence of treatment-risk interactions for relative effects, but moderate risk magnification | Despite a strong overall benefit of corticosteroids, the risk model identified 36% of low-risk patients with almost no predicted benefit (and increased risks corticosteroid-related risks for re-hospitalization and hyperglycemia) |
| Reference | Clinical | Outcome(s) | Randomized | Overall RCT | Methods | Type of HTE | Clinically Important Subgroup |

|  | Condition |  | Interventions | Findings |  | Identified | Differences |
| --- | --- | --- | --- | --- | --- | --- | --- |
| **EFFECT MODELS (n=9)** |  |  |  |  |  |  |  |
| Park et al.<sup>51</sup> | Hospitalized Covid Infection without mechanical ventilation | Ordinal COVID-19 clinical status scale | Convalescent Plasma (CCP) vs control | Null (no overall association between CCP and patient outcomes) | IPDMA of 8 trials; proportional odds model; external validation in multiple datasets. | Relative treatment effect differences demonstrated with non-overlapping 95% confidence intervals | Despite null findings in overall trial population, predictive modeling identified 3 roughly equal-sized subgroups, one with high benefit from CCP, one with modest benefit from CCP, and the third with modest harm from CCP. |
| Takahashi et al.<sup>52</sup> | De-novo three-vessel and left main coronary artery disease | 5-year major adverse cardiac events (MACE) and 10-year all cause death | Coronary artery bypass graft (CABG) vs percutaneous coronary intervention (PCI) | Strong benefit in favor of CABG (HR=0.84 for 10-year mortality; HR=0.78 for 5-year adverse cardiac events) | Single RCT-reanalysis; Cox proportional hazards regression; external validation in IPDMA of 3 trials | Multiplicative interactions of treatment choice with two pre-specified effect modifiers. | Predictive model identifies half of patients who clearly benefit from CABG (vs PCI); and half in whom there is no expectation of greater benefit with the more invasive procedure for either 5-year outcome (MACE) or 10-year all cause death. |
| Dennis et al.<sup>70</sup> | Type 2 diabetes | Hb A1c at 6 mos post-initiation | Initiation of SGLT-2 inhibitors vs DPP-4 inhibitor as add-on therapy | Weak benefit in favor of SGLT-2 inhibitor: mean difference in Hb A1c: 1.2 mmol/mol | Model built in large obs. cohort; linear regression; external validation in multiple RCTs. | Clear subgroup heterogeneity in absolute differences in treatment effects on Hb A1c control. | Predictive model identifies a large subgroup, comprising 60% of trial participants, with strong benefit if SGLT-2 inhibitor is initiated (vs DPP-4 inhibitor); a small group (~5%) appear to do better if a DPP-4 inhibitor is initiated. |
| Seitz et al.<sup>80</sup> | Respiratory Distress | Successful intubation on the first attempt | Bougie vs Stylet | Null (non-significant 6.8 percent point difference in favor of stylet) | Single RCT re-analysis; causal forest; external validation in non-random subset | Absolute and relative effect differences (p=0.02 for multiplicative interaction of treatment with predicted treatment effect). | Despite null findings in the overall trial population, predictive modeling identifies a quarter of patients in the validation cohort who clearly benefit from use of the stylet (vs bougie); a quarter of patients with a possible marginally better result when bougie is used; with no treatment effect differences in the remainder. |
| **Reference** | **Clinical Condition** | **Outcome(s)** | **Randomized Interventions** | **Overall RCT Findings** | **Methods** | **Type of HTE Identified** | **Clinically Important Subgroup Differences** |
| Venkatasu-bramaniam et al.<sup>92</sup> | Diabetes, type 2 | Hb A1c reduction at 6 mos. | SGLT-2 inhibitors vs DPP-4 inhibitors as add-on therapy | A very modest benefit of SGLT-2 inhibitors (1.6 mmol/mol greater Hb A1c reduction at 6 mos) | IPDMA of 2 trials; penalized linear regression and causal forest models, external validation in large observational cohort (n=18,741) | Both penalized regression and causal forest models indicated significant HTE (p=0.005 for causal forest); significance testing not reported for regression model, but benefit prediction in external cohort was excellent | Regression model reliably identified a subgroup of 87% of validation cohort who did better if initiated on an SGLT-2 inhibitor, including 30% of all patients who were predicted to have a >5 mmol/mol larger reduction in Hb A1c if started on the SGLT-2 inhibitor. For 13% of patients, the Hb A1c reduction was predicted to be better with DPP-4 inhibitors. |
| Buell et al.<sup>93</sup> | Patients requiring mechanical ventilation | 28-day mortality | Lower vs higher O2 saturation targets | Null findings in multiple trials of moderate size. | Single RCT re-analysis; ensemble machine-learning effect models (RBoost); external validation in a second RCT | Both relative and absolute HTE (p=0.02 for interaction) shown across tertiles of predicted benefit in the validation trial | Null overall findings in 7 prior RCTs comparing lower vs higher O2 saturation targets trials; these validated analyses identified subgroups of patients who clearly benefited from either lower (33% of all patients) or higher O2 saturation targets and also identified the most critical individual characteristics for predicting benefit. |
| Arnold et al.<sup>99</sup> | Patients with chronic coronary artery disease | Angina symptoms at 12 months follow-up | Coronary revascularization vs medical therapy | A modest benefit of revascularization for angina symptoms | Single RCT re-analysis; ordinal proportional odds regression effect model | Strong absolute HTE, (<0.01) for a single baseline covariate – symptom severity. | Weak overall benefit of revascularization; effect model identified a single strong effect modifier and subgroups of 75% in whom the predicted benefit was strong, and 25% predicted to experience little if any benefit. |
| Ninomiya et al.<sup>108</sup> | Patients with de-novo three-vessel or left main coronary artery disease 3-vessel | 5-year major adverse cardiac events (MACE) and 10-year all cause death | Coronary artery bypass graft (CABG) vs percutaneous coronary intervention (PCI) | Strong benefit in favor of CABG (HR=0.84 for 10-year mortality; HR=0.78 for 5-year adverse cardiac events) | Single RCT re-analysis; proportional hazards regression effect model with two pre-specified interactions; external validation in observational cohort | Strong absolute and relative scale HTE (p<0.005 for two covariates) | Despite relatively strong overall benefit of CABG, effect model identifies 25% of patients who would have better predicted outcomes with PCI, 50% with very strong benefit of CABG, and 25% for whom there is little predicted difference in benefits. |
| **Reference** | **Clinical Condition** | **Outcome(s)** | **Randomized Intervention** | **Overall RCT Findings** | **Methods** | **Type of HTE Identified** | **Clinically Important Subgroup Differences** |
| Smit et | Community- | 30-day | Corticosteroids | Strong benefit | IPDMA of 6 trials; | Strong HTE driven | Despite a relatively strong overall |
| al.<sup>111</sup> | acquired pneumonia | mortality | vs placebo | of corticosteroids (OR 0.72; 95% CI 0.56-0.94) | penalized logistic-regression based effect models; external validation in 2 addition RCTs. | primarily by interaction with a single baseline covariate, the C-reactive protein level. (p=0.05) | benefit of corticosteroids, the effect model identified nearly half of patients, with lower levels of CRP, with no predicted benefit from corticosteroids, and experienced adverse effects including re-hospitalization and hyperglycemia |

## Discussion

Evidence-based medicine has historically encouraged clinicians and patients to rely on average treatment effects from RCTs to support individual decision-making, (23,115) despite recognizing limitations with this approach. In the four and a half years following publication of the PATH Statement, a steadily growing number of publications across a wide range of clinical areas has employed predictive modeling to examine possible HTE in RCT results. Among the 65 reports we identified, more than a third found HTE that was both credible and clinically important, suggesting that patients and clinicians could often do better than relying solely on average effects.

Consistent with the PATH Statement rationale and ICEMAN criteria, risk modeling was more likely than effect modeling to produce findings of credible HTE because of its relative simplicity. Importantly, HTE was not always confined to the absolute scale (risk magnification). In nine reports, credible and important HTE was identified for relative as well as absolute treatment effects across baseline risk. In five, (49,54,61,79,96) relative as well as absolute effects were greater for persons at higher predicted risk. In two, (59,84) persons at low risk benefited while high-risk patients may have been harmed by the same treatment; and in two, (47,69) maximal benefit was found for those in the mid-range of risk. This U-shaped, or “sweet spot,” pattern (47) is clinically intuitive and has also been observed elsewhere. (116) These findings demonstrate that simple assumptions of risk magnification are not well-founded and illustrate the potential value of routine risk modeling of RCT results.

They may offer clinical insights about specific effect modifiers. Traits incorporated into risk scores because they are strong predictors of study outcomes may sometimes also be treatment effect modifiers, either directly or as proxies for unmeasured attributes. In an RCT comparing therapeutic-dose heparin with usual thromboprophylaxis for patients hospitalized with COVID-19, (84) respiratory status at baseline was the most potent predictor of clinical outcomes but was also found to be a strong modifier of heparin treatment effect in initial subgroup analyses. Only patients with better baseline respiratory status benefited from heparin. When this trait was incorporated into a risk model, only patients with lower risk scores appeared to benefit. In three RCTs (47,59,69) where incidence of study outcomes was particularly high (range 27-61%), no benefit was observed in the highest stratum of predicted risk. For such extremely high-risk individuals, risk prediction models likely included attributes reflecting irreversible disease or competing causes of the outcome that would make treatment futile.

The increasing use of effect modeling, particularly exploratory machine-learning approaches, suggests enthusiasm for moving beyond risk stratification to more flexible estimation of individualized treatment effects. Several authors (58,76,88) expressed concerns that although risk scores can create patient subgroups well-matched on risk, subgroup members may still be heterogenous for the specific characteristics that contributed to their risk scores and therefore potentially heterogeneous in their responses to treatment. Comparative advantages and disadvantages of risk versus effect modeling remain incompletely understood. Seven reports (57,81,83,91,105, 109,111) presented both risk and effect modeling of the same RCT data. Effect modeling added new insights to risk modeling in only one instance. (111) In this IPDMA of eight RCTs of corticosteroids for community-acquired pneumonia, effect modeling identified a single powerful relative treatment effect modifier, c-reactive protein, a variable that was not included in the external risk model. Although both models found credible HTE, the effect model performed better in external validation. More generally, the existence of strong effect modifiers, whether known in advance or not, is a likely pre-requisite for finding that an effect model improves on risk modeling.

Nearly all effect modeling reports followed Statement suggestions to use shrinkage methods and internal validation strategies to reduce over-fitting (Supplement Table 4). Nevertheless, inconsistencies in several reports illustrate the persistent challenges of false positive signals of HTE in exploratory effect modeling and underscore the need for external validation. For example, two reports (55,82) applied causal forest algorithms to data from the SPRINT and Action to Control Cardiovascular Risk in Diabetes (ACCORD) trials evaluating intensive systolic blood pressure control. One (82) found evidence of HTE, the other did not. In two reports (60,78) from a trial of dabigatran vs. warfarin for stroke prevention in atrial fibrillation, one (60) suggested interactions of three covariates with treatment choice and significant HTE; the second, using four machine-learning algorithms applied to the same RCT data, found no evidence for HTE. Two reports (62,110) compared results of multiple machine-learning algorithms, finding inconsistent evidence for HTE between algorithms and even within algorithms when random initiation seeds were altered.(62) Two reports (92,111) compared regression-based methods with machine-learning algorithms in effect modeling, both finding that regression models performed better in external validations.

The inconsistency of machine-learning approaches to effect modeling has also been addressed by others (117). Most effect models reviewed here explored large numbers of candidate treatment interactions despite relatively modest numbers of outcome events. Specification of best practices in this emerging area is beyond the scope of the present review, and much remains to be learned concerning sample size needs and optimal approaches to internal validation.

Meanwhile, the value of external validation illustrated here highlights the importance of making data from completed RCTs available and of creating large, well-characterized real-world cohorts with treatment and covariate data for validation and extending HTE findings to populations with differing patterns of risk and treatment. (118) The same cohorts could support development of new, more representative risk prediction models.

Given the frequent utility of risk modeling, sponsors of new RCTs should consider in advance whether appropriate external risk models exist and plan for collection of baseline data needed for estimating individual risk. Within the past decade, editorial guidelines for reporting positive RCTs findings have come to require presentation of absolute as well as relative measures of overall treatment effect because of their greater relevance in clinical decision-making. (119–121) We suggest that reports of positive RCTs could be further enhanced by requiring that treatment effects, in both relative and absolute terms, be presented in relation to baseline risk. Ultimately, it will remain critical to demonstrate the safety and effectiveness of any predictive model when employed for personalizing treatment choices in real world populations.

## Limitations

The search strategy would not have captured predictive modeling reports that did not cite the PATH Statement. We conducted a broad search for such reports using title/abstract words “randomized” plus “heterogeneity of treatment effects” for the same time period. This strategy yielded 60 publications, but only seven were predictive models in RCTs, 3 of which were included in our review. That this search found so few of the 65 reports we reviewed indicates that it is not a parallel approach for finding other predictive models. Nevertheless, we believe that the reports citing the PATH Statement offer a highly relevant population for assessing its influence. Even with the ICEMAN and PATH criteria for assessing credibility and clinical importance of HTE, some subjectivity remains. The close association of two authors (DK, JS) with production of the PATH Statement should be kept in mind.

## Conclusions

The PATH Statement appears to be influencing research practice. Although effect modeling holds promise for predicting individualized treatment effects, the need for external validation is a constraint. Risk modeling provides a more straightforward initial approach when overall trial findings are positive and often identifies clinically important HTE.

## Data Availability

All data produced in the present study are available upon reasonable request to the authors

## Acknowledgments

The authors gratefully acknowledge Harold Sox, MD, Department of Medicine and The Dartmouth Institute (emeritus), Geisel School of Medicine at Dartmouth, Hanover, NH, for careful review and helpful suggestions on earlier drafts of the manuscript; Jinny G. Park, MPH, Tufts Predictive Analytics and Comparative Effectiveness Center, Tufts University School of Medicine, Boston, MA, for conducting all literature database searches; and Ivan Rivera, MIS, Division of Research, Kaiser Permanente Northern CA, for retrieving reprints and supplemental materials of study citations.

