## Supplemental materials for "Potential clinical impact of predictive modeling of heterogeneous treatment effects: scoping review of the impact of the PATH Statement"

**Potential clinical impact of predictive modeling of heterogeneous treatment effects: scoping review of clinical trials analyses that cite the PATH Statement**

**SUPPLEMENTARY MATERIAL**

**Jan 5, 2025**

Joe V Selby

Carolien C H M Maas

Bruce H Fireman

David M Kent

[S7. Instructions for applying ICEMAN Criteria to claims of HTE	2](#_Toc171066256)6

S1. Search and Inclusion/Exclusion Criteria

**Introduction**

Eligible reports for this scoping review were identified using the “Cited By” functions in four search engines: PubMed, Google Scholar, Web of Science and the SCOPUS database to find reports appearing after publication of [The Predictive Approaches to Treatment Effect Heterogeneity (PATH) Statement.](https://pubmed.ncbi.nlm.nih.gov/31711134/) (January 7, 2020 through June 5, 2023) that cited the Statement, presented analyses or re-analyses of data from one or more RCTs and that used multivariable predictive modeling to identify HTE. Table 1a below presents the search terms. Restricting the search to articles citing the Statement allows the assumption that authors were aware of its concepts and recommendations. Including sources other than PubMed allowed inclusion of non-peer-reviewed reports posted on pre-print archives as well as dissertations posted on institutional websites.

The PATH Statement defined predictive modeling as multivariable modeling intended to provide patient-centered [i.e., individualized] estimates of outcome risks with versus without the intervention, taking into account all relevant patient characteristics simultaneously. Not all of the studies that cited the Statement and analyzed RCT data met this definition. Some performed one-at-a-time tests for a variety of potential effect modifiers. Others focused on hypotheses about one or two potential effect modifiers. Some of these papers included multivariable models used to adjust for potential confounding between subgroups, but the focus remained on hypothesis-testing, one variable at a time.

Table 1b below identifies thirteen reports that were identified by our initial search and found to have analyzed RCT data, but judged to not be a predictive model as defined in the PATH Statement. In nine cases, authors were focused on understanding potential effect modification by one or two patient variables; in three reports, one-at-a-time analyses of multiple patient characteristics were presented but no attempt was made to include all in a single predictive model. In the final report (Inoue et al), a multivariable approach was used to generalize counterfactual findings from an RCT population to various hypothetical populations with different baseline demographic and clinical characteristics.

#### Supplement Table 1A. Electronic Search Strategy

Citations of (‘The Predictive Approaches to Treatment effect Heterogeneity (PATH) Statement’ OR ‘The Predictive Approaches to Treatment effect Heterogeneity (PATH) Statement: Explanation and Elaboration’) AND [2020-2024]/py

#### Supplement Table 1B. 16 Reports Excluded from Review Because They Do Not Meet PATH Definition of a Predictive Model

| **Reference^*^** | **Clinical Condition** | **Outcome** | **Randomized Intervention(s)** | **Approach** |
| --- | --- | --- | --- | --- |
| Kataoka et al^30^ | Hyperuricemia in kidney failure | Progression of chronic kidney disease (estimated glomerular filtration rate) | Febuxostat vs placebo | Focus is exclusively on possible interactions of baseline proteinuria and serum creatinine level with effectiveness of urate-lowering therapy with febuxostat. |
| Albuquerque et al^31^ | Patients hospitalized with COVID-19 | All-cause mortality; hospital discharge within 28 days | Tocilizumab vs placebo | Focus is on possible interactions of concurrent corticosteroid use and baseline ventilatory support requirements with effectiveness of tocilizumab. |
| Foy et al^32^ | Hypertension | All-cause mortality; composite cardiovascular outcome | Intensive vs. standard systolic blood pressure control | Focus is exclusively on the single possible interaction of initial diastolic blood pressure with effectiveness of intensive systolic blood pressure treatment. |
| Kloecker et al^33^ | Type 2 Diabetes | Composite cardiovascular outcome | Intensive vs usual glycemic control | Focus is exclusively on the single possible interaction of diabetic microvascular disease at baseline with effectiveness of intensive glycemic control. |
| Dianti et al^34^ | Acute Respiratory Failure | 90-day mortality | Extracorporeal CO_2_ removal (ECCO_2_R) | Focus is exclusively on possible interactions of ventilatory ratio and respiratory system elastance with the effectiveness of ECCO_2_R. |
| Farrar et al^35^ | Interstitial Cystitis, Bladder Pain | Change in pain scores | Multiple treatments | Focus is exclusively on the single possible interaction of the extent of widespread pain on the effectiveness of several pain treatments. |
| Hanlon et al^36^ | Multiple diseases | Various outcomes in 120 industry sponsored Phase III/IV trials | Various treatments | Focus is exclusively on the single possible interaction of comorbidity measured in a variety of ways with treatment effectiveness across 120 trials. |
| Samuels et al^37^ | Acute ischemic stroke | Functional Outcome (Modified Rankin Scale) at 90 days | Endovascular thrombectomy vs medical therapy | Focus is exclusively on the single possible interaction of admission systolic blood pressure with the effectiveness of thrombectomy vs medical therapy. |
| Kimchi et al^38^ | Heart failure | 30- and 180-days mortality, readmission, days alive and out of hospital | Nurse Tele-monitoring and telephone coaching vs. usual care | Focus is on the single possible interaction of level of comorbidity with the effectiveness of nurse telemonitoring and telephone coaching. |
| Gargiulo et el^39^ | Acute coronary syndrome | All-cause mortality; major bleeding | Radial vs femoral artery access | One-at-a-time exploratory assessment of multiple patient characteristics as possible effect modifiers of the superiority of trans-radial vs transfemoral percutaneous arterial access. |
| Klitgaard et al^40^ | Respiratory failure | 90-day all-cause mortality | Lower vs higher inspired oxygen | One-at-a-time exploratory assessment of four patient characteristics as possible effect modifiers of the impact of lower vs. higher oxygen levels. |
| Wijn et al^41^ | Degenerative  meniscal tear of knee | Pain, function and quality of life at 24 months | Arthroscopic partial meniscectomy vs. non-surgical or sham treatment | One-at-a-time exploratory assessment of 10 patient characteristics for possible interaction with the relative effectiveness of partial meniscectomy in patients with degenerative meniscal disease. |
| Inoue et al^42^ | Lung Cancer | Lung Cancer mortality | Computed tomographic (CT) screening | Examines the generalizability and transportability of findings from the National Lung Screening Trial to other populations using inverse odds weighting |
| Foy et al^43^ | Various cardiovascular illnesses | Various cardiovascular disease outcomes | Various interventions in 8 RCTs | Focus is exclusively on the single possible interaction of chronic disease comorbidity on the effectiveness of cardiovascular disease interventions in 8 RCTs |
| Bertsimas et al^44^ | Sarcomas of the extremities | Extremity or truncal sarcomas | Prophylactic radiation therapy | Uses predictive modeling solely to subgroup patients on risk of cancer recurrence. Does not examine treatment effectiveness, either overall or in subgroups. |
| Cheng et al^45^ | Employed adults | Weight change, dietary intake | Employment-based lifestyle intervention | Focus is exclusively on the single possible interaction of the presence of a chronic illness on the effectiveness of a lifestyle intervention. |
| ^*^ Reference numbers refer to the bibliography in the manuscript. | | | | |

### S2. PATH Statement Consensus Criteria

The boxes in Table S2 below are taken from the PATH Statement. For the second criterion in Box A (risk modeling), we did not feel that reviewers could objectively evaluate whether or how well researchers had “integrated clinical and statistical reasoning and expertise” in conducting and reporting their analyses, although some authors clearly attempted to do so more than others. The tenth criterion is not a clear suggestion. Rather, it is simply a description of a possible approach that could be taken, without a value statement attached. Therefore, only 8 of the ten criteria were assessed.

#### Supplement Table 2. PATH Statement Consensus Criteria

**Box A. Risk Modeling Approaches to Identify HTE***

**General**

1. Reporting RCT results stratified by a risk model is encouraged when overall trial results are positive to better understand the distribution of effects across the trial population.
2. Predictive approaches to HTE require close integration of clinical and statistical reasoning and expertise.^†^

**Identify or Develop a Model**

3. When available, apply a high-quality, externally-developed, compatible risk model to stratify trial results.
4. When a high-quality, externally-developed model is unavailable, consider developing a model using the entire trial population to stratify trial results; avoid modeling on the control arm only.
5. When developing new risk models or updating externally-developed risk models, pre-specify the analytic plan prior to examination of trial data and follow guidance for best practice for prediction model development.

**Apply the Model and Report Results**

6. Report metrics for model performance for outcome prediction on the RCT, including measures of discrimination and calibration (when appropriate).
7. Report distribution of predicted risk (or the risk score) in each arm of the trial, and in the overall study population.
8. Report outcome rates and both relative and absolute risk reduction across risk strata.
9. When there are important treatment-related harms, these harms should be reported in each risk stratum to support strata-specific evaluation of benefit-harm trade-offs.
10. To test the consistency of the relative treatment effect across prognostic risk, a continuous measure of risk (e.g., the logit of risk) may be used in an interaction term with treatment group indicator.^‡^

**Box B. Considerations and Caveats in Effect Modeling for HTE***

1. When highly credible relative effect modifiers have been identified, they should be incorporated into prediction models using multiplicative treatment-by-covariate interaction terms.
2. Credibility should be evaluated using rigorous multidimensional criteria (such as described in the ICEMAN tool) and should not rely solely on statistical criteria (such as p-value thresholds).
3. Avoid one-variable-at-a-time null hypothesis testing or stepwise selection (e.g., backward selection, forward selection) strategies to select single variable relative effect modifiers.
4. Avoid the use of regression methods that do not take into account model complexity when estimating coefficients (e.g., “conventional” unpenalized maximum-likelihood regression) when one or more treatment by covariate interaction terms are included in a treatment effect model.
5. Avoid evaluating models that predict treatment benefit using only conventional metrics for outcome risk prediction (e.g., metrics based on discrimination and calibration of outcome risk prediction).

* Adapted from: Kent DM, Paulus JK, van Klaveren D, D'Agostino R, Goodman S, Hayward R, Ioannidis JPA, Patrick-Lake B, Morton S, Pencina M, Raman G, Ross JS, Selker HP, Varadhan R, Vickers A, Wong JB, Steyerberg EW. [The Predictive Approaches to Treatment effect Heterogeneity (PATH) Statement.](https://pubmed.ncbi.nlm.nih.gov/31711134/) Ann Intern Med. 2020;172:35-45.

^†^ Determination of this guidance statement was not within the scope of this review.

^‡^ This guidance statement was not considered to be a clear recommendation.

### S3. Variables Collected, Possible Values and Initial Disagreement Rates

#### Supplement Table 3. Variables Collected, Possible Values and Initial Disagreement Rates for Items Doubly Reviewed

| **Variable** | **Possible Values** | **Initial Disagreements/ Number of Assessments (%)*** |
| --- | --- | --- |
| ***Variables collected by single review*** | | |
| Year of Publication or Posting | 2020-2023 |  |
| Most recent Publication Status | 1=Published; 2=Preprint archive; 3=Posted dissertation |  |
| Nature of Report | 1=Re-analysis of single RCT, 2=IPDMA of ≥ 2 RCTs, 3=Initial report of RCT(s) |  |
| Initial Trial Results | 0=Neg, null; 1=Weak benefit; 2=Strong benefit |  |
| Which treatment effect scale(s) are presented? | 1=Relative scale only; 2=Absolute scale only; 3=Both scales |  |
| Is this a comparative effectiveness study? | 0=No; 1=Yes |  |
| Is there a need to balance between harms/benefits? | 0=No; 1=Yes |  |
| Is there risk of serious adverse effects with ≥1 treatment? | 0=No; 1=Yes |  |
| Are critical variables available in clinical care? | 0=No; 1=Yes |  |
| ***Variables collected by double review*** | | |
| Does the report present a predictive model? | 0=No; 1=Yes | 2/65* |
| Does the report present a risk model? | 0=No; 1=Yes | 4/65* |
| Does the report present an effect model? | 0=No; 1=Yes | 5/65* |
| ***For Risk Models Only (n = 28*)*** | | |
| Was the risk model developed externally or internally to the RCT(s) | 1=external, 2=internal | 2/29 |
| If internal model is used, in what sample is it developed? | 1=Entire sample; 2=Controls only | 2/15 |
| Were risk modelling analyses pre-specified? | 0=No; 1=Yes | 4/29 |
| Were performance metrics of the risk model in the RCT population reported? | 0=No; 1=Yes | 1/29 |
| Was the risk score predicted risk reported by arm? | 0=No; 1=Yes | 2/29 |
| Were important harms reported by stratum? | 0=No; 1=Yes | 1/29 |
| Is a multiplicative treatment by baseline risk interaction inferred by authors? | 0=No; 1=Yes | 2/29 |
| Is a statistical test for multiplicative interaction presented? | 0=No; 1=Yes, p<0.05; 2=Yes, p≥0.05 | 3/29 |
| Reviewer assessment of risk magnification or multiplicative interaction? | 1=Multiplicative interaction; 2=Risk magnification; 3=Neither/unclear | 3/29 |
| ***For Effect Models Only (n = 36*)*** | | |
| Type of effect model? | Regression-based model; 2=More flexible machine-learning model | 2/36 |
| Was validation in an external dataset performed? | 0=No; 1=Yes | 1/36 |
| ***Assessment of Credibility and Clinical Importance of HTE (n=54)*** *^§^* | | |
| Did the authors test only a small number of potential effect modifiers? (ICEMAN #1) | 1=Definitely not; 2=Probably not; 3= Probably; 4=Definitely | 5/54 |
| Does each effect modifier have support from prior evidence of possible effect modification? (ICEMAN #2) | 1=Prior evidence suggests effect modification in a different direction; 2=No prior evidence/unclear; 3=Some support; 4= Strong support | 5/54 |
| Are any cut-points for continuous effect modifiers pre-specified? (ICEMAN #3) | 1=Cut-points derived from the RCT data; 2=Unclear; 3=Cut-points suggested from prior RCT analyses; 4=Effect modifiers analyzed as continuous variables | 12/54 |
| Does a test for interaction suggest that chance is an unlikely explanation of the apparent effect modification? (ICEMAN #4) | 1= Interaction p-value > 0.05; 2= Interaction p-value ≤ 0.05 but > 0.01, or no test of interaction reported and not computable; 3= Inter-  action p-value ≤ 0.01 and > 0.005; 4=Interaction p-value ≤ 0.005. | 7/54 |
| Credible HTE? | 0=No; 1=Yes | 11/54 |
| Reviewer Assessment of Clinically Important THE? | 0=No; 1=Yes | 10/54 |
| Overall Disagreement Rate | 84/838 = 10.1% | |
| * After removing six reports reviewed in the initial training exercise (covering 6 effect models and one risk model), there were 59 reports doubly reviewed, presenting 65 predictive models (29 risk models, 36 effect models. Six reports presented both a risk model and an effect model.  ^§^ Credibility of HTE and clinical importance of claimed HTE was assessed in the 54 reports in which authors claimed to have found HTE on either the absolute or relative scale (or both). | | |

### S4. Review Coding Instructions

#### Supplement Table 4. PATH Follow-up Study Coding Instructions

**July 17, 2023**

| A. **Variables to be Recorded By Single Reviewer (JS)** | | | | |
| --- | --- | --- | --- | --- |
| **#** | **Variables** | | **Instructions** | |
| S1 | Publication Year | | Enter year of publication for all peer-reviewed publications. For those found in pre-print archives or online dissertations, enter date associated with appearance on the archive, or completion date shown on the dissertation itself**.** If an article in a pre-print archive is subsequently published in a peer-reviewed publication and found on Pub Med, change this date to the publication date given on Pub Med | |
| S2 | Publication Type  (1= peer-reviewed; 2=preprint archive; 3=dissertation) | | If the article has been published, regardless of whether it was first discovered via PubMed or Google Scholar, list this as a peer-reviewed article. Otherwise, if it is found via Google Scholar only on a pre-print archive, or as a posted dissertation, list it as such. | |
| S3 | Link | | For peer-reviewed articles, enter URL of PubMed abstract; for articles on preprint archives, enter URL for article on archive website; for dissertations, enter URL at which dissertation was accessed. If the article is initially posted as a pre-print and subsequently published, replace the link with the PubMed link or other link to the published article. | |
| S4 | Type of Analysis – (IPDMA;  RCT-reanalysis;  RCT initial analysis) | | List as an IPMDA if data from two or more trials are analyzed and reported – whether data are actually pooled prior to analyses or reported as similar parallel or stratified analyses. It is the act of using individual data from multiple trials that is of most interest; list as an RCT re-analysis if data from a single trial are analyzed AND overall results from this same trial have been previously published; list as initial analyses if the analyses for treatment heterogeneity are reported in the same article that is reporting overall treatment effect for the first time. | |
| S5 | First Author | | Record last name of first author | |
| S6 | PATH Co-author  (0=NO, 1=YES) | | Examine author list of each paper to determine whether any co-author was also listed among the authors of the PATH Statement. | |
| S7 | Disease | | Describe disease or condition under study, whether the study concerns treating the disease, screening for it, or preventing it. If a treatment is used in a baseline condition to prevent a second condition (e.g., anti-coagulant to prevent pulmonary embolism (PE) in patients with cancer), enter the baseline condition (e.g., cancer) not the outcome (i.e., PE). | |
| S8 | Treatment | | Using prose, enter the treatment or treatments being studied. In the case of two or more treatments being compared for the same condition, use “vs” to express the comparison. | |
| S9 | Initial Trial Results 0=neg, null; 1=weak benefit 2=strong benefit 3=not available/unclear | | This refers to the interpretation of the authors of the overall, or average, treatment effect (usually published previously) at the time they undertook studies of possible heterogeneity of treatment effect. This may be based solely on the study(ies) being re-analyzed or it may represent a broader consensus based on multiple prior studies. For articles reporting the initial results of a trial with no pre-existing consensus (e.g., in treatments for Covid-19), look to the overall effect reported in the same paper and use that effect to score this variable.  If no effect has been demonstrated (null study), or if the consensus appears to be that an effect has not been established, Score = 0). If an effect has been established, and it is modest (less than a 20% reduction in relative risk associated with one treatment compared either to no treatment, a placebo, or an alternative treatment), Score = 1. If a stronger effect (≥ 20% reduction in relative risk, compared to either no treatment, a placebo, or an alternative treatment), Score = 2. For continuous outcomes, a standardized mean difference significantly >0­, but ≤ 0.8 should be scored as modest (=1); and a standardized mean difference >0.8 should be scored as strong (=2). In the rare situation in which it is unclear whether the trial being re-analyzed shows an overall effect and there is no consensus from other studies, Score = 3. | |
| S10 | Scale(s) Used 1=Relative only  2=Absolute only  3=BOTH | | H.T.E. is often assessed only on the relative scale (OR, HR, RR), especially in effect modelling approach. However, in some papers, only the absolute scale (i.e., risk difference, survival difference, or difference in other continuous outcome such as HbA1c) is examined. The PATH statement encourages examination on both scales, but specifically encourages presenting results in terms of absolute benefit – that being more useful to clinicians and patients for individual decision-making. To score, give credit for “both” only if both scales are presented throughout the results. Otherwise, if one predominates heavily, score only that one. Single mentions of a HR or alternatively of a RD, should not cause you to score “both”. | |
| S11 | Disease vs Primary Prevention 1=Disease, 2=Primary prevention, 3=non-health | | Most studies will be related to treatment for a specific condition (1) since we are studying treatment heterogeneity. However, the same concept (that the intervention may work differently as a function of subject characteristics) can apply to primary prevention and screening interventions (2) in healthy populations, and also to non-medical interventions, such as interventions in education or the legal system (3). | |
| S12 | CER? 0=no, 1=yes | | CER is the comparison of two or more ACTIVE approaches to treating a condition and/or preventing an adverse outcome. For our purpose, comparisons of an active intervention to "usual care" would not be considered CER if both arms got the usual care and only one arm received an additional “new” treatment; but would be considered CER if one arm received usual care (either newly assigned or ongoing) while the other arm received the “new” or alternative treatment instead. | |
| S13 | Need to balance between harms/benefits? 0=NO, 1=YES | | Answer yes if this is discussed by authors or is known or obvious. This is the situation when there are both expected benefits and known adverse effects – but none are over-riding and patients may differ in their preferences and/or other risk aversion, leading to heterogeneity in benefit-risk calculations such that information on differential effectiveness by risk level would be highly valuable in making the individual decision | |
| S14 | Risk of serious adverse effects with ≥1 treatment? 0=NO, 1=YES | | Here we are looking for one or more adverse effects that anyone would like to avoid, especially if they learned that the treatment actually had little or no effectiveness for them. That is, the adverse effect is an over-riding concern of patients and clinicians considering this treatment. | |
| S15 | Critical variables available in clinical care?  0=NO, 1=YES | | Score this as a “1” for any study that presents a risk prediction model **if** the model components are available in clinical care. This may include variables that are routinely collected as well as variables that could easily be collected, but not complex diagnostics not currently used for clinical purposes. Also score as a “1” if the authors use an external risk prediction model are all of its components are available. Otherwise, score as “0”. | |
| B. **Variables to be Collected by Two Reviewers, Compared and Adjudicated** | | | | |
| **#** | | **Variable** | | **Instructions** |
| **General Questions** | | | | |
| D1 | | Does this Paper present a Predictive Model? (0=NO, 1=YES) | | From the PATH Statement: “The goal of predictive HTE analysis is to provide individualized predictions of treatment effect, specifically defined by the difference between expected potential outcome(s) of interest in a particular patient with one intervention versus an alternative—taking into account multiple relevant characteristics simultaneously.” Note that one-at-a-time analyses of possible effect modifiers are NOT a predictive model, even if they are multiply adjusted. Risk models as well as multivariable effect models that generate cATEs (or ITEs) are considered predictive models. Importantly, **authors may construct and report predictive models without the explicit intention that it be used to predict individual treatment effects –** i.e., they may be using the model to explore possible HTE or be examining the model or models for methodologic purposes. Score this as “1” if there is a predictive model, regardless of author’s intent. Otherwise, score “0”. |
| D2 | | Notes on Predictive Model | | Explain your thinking, especially if you do NOT consider this a predictive model |
| **If D1 = 0; stop here.** | | | | |
| D3 | | If yes, do authors provide information necessary to implement the model? (0=NO; 1=yes, in a ready-to-use calculator online or in paper; 2=coefficients or regression tree results and visual graphics allow individual risk calculation and absolute risk reduction prediction) | | In some papers, the authors provide a readily implementable tool, either in the paper or via the supplement or an on-line calculator, to implement their model’s predicted treatment effect for an individual patient. Score such papers as a “1”. In others, there is already a published and readily implementable external prediction model – usually a risk prediction model – that can be combined with treatment effect data in the paper based on this model. Either of these would also be classified as “1”. In other papers, the coefficients from the prediction model(s) are provided, along with an intercept or some other way of ascertaining the “baseline” value of the outcome (could be a continuous endpoint, an odds or a hazard), so that each individual’s predicted absolute value for the outcome can be calculated based on coefficients for the relevant covariates. If what is presented meets this criterion, classify the paper as “2”. Otherwise, If the paper doesn’t provide sufficient information to allow such individual calculations, classify it as “0”. |
| D4 | | Notes on Information Provided to Implement model | | Explain your thinking. |
| D5 | | Authors' Primary Aim: 1=Pred Model 2=Explore/Identify HTE 3=Evaluate HTE Methods 4=Other | | Although you’ve decided that the authors are reporting a predictive model, not all authors express the primary aim of producing a model intended to predict individual treatment effects. If they do state their intention to produce a model for this purpose, score as “1”. Some are explicit that they are “exploring” for possible HTE and others may be seeking to confirm specific hypotheses about possible effect modifiers but using predictive models to do so; score either of these two groups as “2”. Some others are explicitly investigating methodological questions about predictive models; score these as “3”. If the primary aim is unclear; score as “4”. |
| D6 | | Does paper report a risk model? (0=NO, 1=YES) | | Risk Modelling studies first construct a multifactorial risk prediction score (either from an external model or score or alternatively developed in the trial population). In some cases, an “ad hoc” risk score may be created based on markers of disease severity and/or comorbid conditions. In all cases, the risk score is applied to each individual’s baseline data, and then the relationship of treatment effectiveness with this baseline risk score is examined. **If answer to this question is “no”, skip questions D8 to D18.** |
| D7 | | Does paper report an effect model? (0=NO; 1=Yes, EM; 2=Yes, Rx Optimization; 3=yes, Unclear) | | In effect modelling, a regression model (or a more flexible Machine learning model) is developed directly on RCT data that includes covariates AND a treatment assignment variable. In many cases, interaction terms of treatment with covariates are included. In others, the models are developed separately in each arm of the trial and “individual treatment effects” are obtained one individual at a time by subtracting predicted outcomes or probabilities of outcomes under one treatment from those under the other (or placebo or no treatment).  Analyses may use traditional regression models (linear, logistic, proportional hazards models, proportional odds) or more flexible machine learning approaches (causal forests; other tree-based methods such as decision trees, recursive partitioning, random forests; or ensemble methods).  The possible existence of interactions and testing for them may be pointed out explicitly or simply included in calculation of individual treatment effects (ITEs) or conditional aggregate treatment effects (cATES).  Treatment regimen optimization approaches (score = “2”) are a type of effect modelling and may use any of the approaches of effect modelling to calculate ITEs, after which the focus is on comparing predicted outcomes for an entire population under treatment assignment based on ITEs (e.g. treat everyone with an ITE suggesting benefit and no one else) vs. other treatment allocation schemes (e.g., everyone, random selection).  **If this is not an effect model, skip questions D21 to D26.** |
| **SKIP QUESTIONS D8 TO D20 IF NOT A RISK MODEL** | | | | |
| D8 | | If a risk model, what type of model? (1=external; 2=internal) | | The The multivariable risk score may come from a model developed and validated previously in other populations (external), from a prediction model that is developed within the clinical trial dataset (internal), or from a more ad hoc measure (e.g., the COVID severity measure or other combinations of clinical variables that are shown to be predictive of outcome). If these ad hoc measures have not been validated previously score their use as an “internal” model. |
| D9 | | If risk model is internal, in which population was it developed? (1=entire trial population, 2=control group only  3=not applicable) | | The PATH Statement recommends that the entire trial population (both control and intervention arm(s) be used in developing an internal prediction model. **Put “not applicable” IF THE RISK PREDICTOR WAS BUILT USING AN EXTERNAL MODEL OR AN AD HOC RISK SCORE.** |
| D10 | | Was the risk modelling plan pre-specified? (0=NO; 1=YES; 2=UNCLEAR) | | Review the abstract and the methods section and any supplemental material to see whether the risk modelling plan (including the approach to stratification of the risk score or parameterization of the score in a model with treatment) was specified before any analyses were completed. If they simply appear in the results section without evidence of pre-planning, so indicate (i.e., score = 0). |
| D11 | | Are performance metrics reported on the risk model? (0=NO; 1=YES) | | This refers to the metrics for either an internally developed riks model or for the fit of an external model to the trial population. Give credit if either discrimination statistics (e.g. a C-statistic, AUC, or simple chi-square for concordance) OR calibration curves/statistics are presented. |
| D12 | | Was the risk model validated? (1=yes, externally; 2=yes, only internally; 3=not validated) | | The PATH Statement emphasizes the importance of external validation for a risk model that is developed internally within the trial population. However, many internally developed models will stop at doing internal validation only. If the authors used an external model, it is safe to consider it “externally validated;” If both internal and external validation were performed on an internally developed model, score as “1” (external validation) because external validation takes priority. |
| D13 | | Is risk score presented by arm of trial? (0=NO, 1=YES) | | This is often presented without much comment, in a table or graphic, or in the supplement. It is a way of examining the success of randomization, the exchangeability of the treatment arm samples, and the lack of confounding. |
| D14 | | Are “significant adverse effect” differences reported within risk strata? | | “Significant adverse effects” are harmful treatment effects that have been mentioned by the authors or that are widely known and appreciated (e.g., bleeding with anti-coagulants). The key question here is whether, in a risk model analysis, such non-therapeutic outcomes as well as the therapeutic outcomes) are shown by treatment arm within strata of the risk score. Financial burden could also be a significant adverse effect in some circumstances. Score this as “1” if any important risk is shown by stratum, even if others are not. |
| D15 | | Was a multiplicative treatment x baseline risk interaction inferred? (0=NO, 1=YES) | | This refers to what the authors appeared to believe or conclude; namely, that the relative treatment effect varies across levels of baseline risk score? This may have been demonstrated either in stratified analyses (quantiles of the risk score) where differences in relative effect are presented in tables or visually in graphic format, or by testing for a multiplicative statistical interaction between the treatment indicator variable and the continuous risk prediction score in a regression model, but statistical testing is not required for this variable. |
| D16 | | Was a statistical test used to evaluate the interaction? (0=NO, 1=YES, p<0.05, 2=YES, p≥0.05) | | Look for mention of a (multiplicative) interaction test between relative treatment effect and the risk score, usually the risk score will be treated as a continuous variable, but it could be analyzed as the stratified variable. This could also be presented as a statistical difference between the fits of models with vs without interaction terms (e.g., likelihood ratio tests, Akaike’s Information Criterion, or by showing that the 95% confidence intervals of effect sizes across strata of the interacting covariate don’t overlap (though that would be rare). |
| D17 | | If no multiplicative interaction inferred, was risk magnification inferred? (0=NO, 1=YES) | | Again, this refers to what the authors said or appeared to think. In the absence of a multiplicative interaction, was an increase in the absolute treatment effect across the range of increasing baseline risk (so called “risk magnification” or “risk magnification”) suggested by authors. Depending on the focus of the analyses and the methods used, absolute risk differences may or may not be shown graphically or in tabular form. If the authors seem unaware of a probable risk magnification, score this as “0” (no) even if you see it. |
| D18 | | In your opinion, was multiplicative interaction or risk magnification observed? (1=multi- interaction; 2=Benefit magnification; 3=neither; 4=both) | | This is the reviewer’s assessment. Base your assessment on both the size and precision of the observed differences in either relative effect (interaction) or absolute effect (risk magnification) across the range of predicted risk. Consider sample size and statistical tests, but your impression should not require a confirmatory statistical test. Note that in overall negative studies, of which there are several, you may see “neither” interaction nor risk magnification. Note that when both absolute and relative treatment effects increase with baseline risk, this should be scored as multiplicative interaction. The absolute treatment effect in high risk individuals is even greater than would be expected based on baseline risk. |
| **SKIP THE REMAINING QUESTIONS IF NO EFFECT MODEL (Q D7=NO)** | | | | |
| D21 | | Type of Prediction Model (1=regress- ion model; 2=more flexible M-L approach) | | In many cases, traditional regression models, OLS, GLM, proportional hazards or logistic regression models are used. Bayesian versions of regression are included here. Although shrinkage methods such as LASSO, elastic net, penalized ridge) are sometimes considered to be forms of machine learning, include these as regression models too if no other machine-learning algorithm is mentioned. Score these as “1”. In others, one or more machine learning algorithms such as causal forests, neural networks, decision trees, or ensemble methods are used to specify the model. Score this as “2”. If both regression and ML methods are used in same paper, score as “2”. |
| D23 | | Were shrinkage or penalization methods used? (0=NO, 1=YES) | | Shrinkage or penalization strategies may be used with either regression-based models or more flexible machine learning approaches. Examples are elastic net estimation, ridge penalization, LASSO methods. In decision trees, restriction on the number of nodes or minimum node size are also a form of shrinkage. Bayesian analyses also employ shrinkage. Shrinkage may be applied to variable selection, estimation of model coefficients, and/or specification and/or identification of interactions. |
| D24 | | Was either internal cross-validation or internal validation on withheld sample used in deriving model estimates? (0=NO; 1=validation performed in the same data used for model derivation to reduce over-fitting; 2= validation on a separate, withheld portion of data not previously included in any aspect of model derivation; 3=both cross-validation and validation in separate portion of data) | | The term internal validation is used for two distinct steps in the derivation and evaluation of a model. Internal cross-validation as well as bootstrapping and other iterative strategies can be used simply to optimize the model fit to the data. If internal cross-validation was used solely for this purpose, or if internal validation is not mentioned, score “0”. However, in the internal assessment and tuning of the original model, averaging model performance over iterative samples (“folds”) from the original data is sometimes used to “correct for optimism” or to “improve generalizability” and is considered a form of shrinkage or regularization. Score these as “1”. If a random portion of the original trial population is set aside, not used in model derivation but only for performance assessment of the final model, score as “2”. If both internal cross-validation and validation in a separate portion of the data, score as a “3.” |
| D25 | | External Validation  (0=NO; 1=YES) | | Did the authors validate the predictive model’s performance in a separate dataset (either another RCT, an external observational cohort, or a non-random subset of the original trial dataset in which the covariates and their estimated effects from the derivation are applied unchanged to the new subset. (0=NO; 1=YES). |
| D27 | | Were treatment by patient characteristic interactions inferred? (0=NO, 1=YES) | | In some analyses, statistical tests may not have been explicitly reported. Only visual differences in treatment effect are shown between patient subgroups. In effect modeling, and especially machine learning algorithms, these are often expressed only as differences in absolute effect sizes. Here we are interested in either situation, as long as the authors showed differences and suggested that they were real and noteworthy. Note that some “treatment optimization” papers do NOT even address specific patient characteristics. They simply suggest that patients have been divided into subgroups with different expected individualized treatment effects. |
| D28 | | In your opinion, was treatment heterogeneity observed? (0=NO; 1=YES; 2=UNCLEAR. | | This is the reviewer’s assessment. Was treatment heterogeneity demonstrated – on either the relative or absolute scale? Take into account absolute treatment effect differences presented, various approaches to testing of statistical significance, and the quality and reliability of the analyses. “Statistical significance” is not essential and will often not be tested, especially in the treatment optimization papers. |
| D29 | | ICEMAN #1: Is a small number of effect modifiers tested? (1=definitely not; 2=probably not; 3= probably; 4=definitely) | | ICEMAN recommends that a very small number of effect modifiers be tested. Consider the original number of variables included in the model or ML algorithm that could appear to be effect modifiers (not the final number that come out after testing many more). Score as 1 if more than 10 effect modifiers were originally considered; 2=if the number is unclear or is between 4 and 10; 3= if the number appears to be 3 or fewer but a protocol is not available to prove this; 4=if the number is 3 or fewer and there is a study protocol that establishes that these were the only ones tested. (Note that pure risk models test one effect modifier – the risk score). |
| D30 | | ICEMAN #2: Does each effect modifier have prior evidence of possible effect modification? 1= PE actually suggests different direction; 2=no PE or unclear; 3= some support; 4= strong support; | | According to ICEMAN, each variable considered and tested for effect modification should have prior evidence. So score this for each variable tested and record the lowest score for any variable:  1=prior evidence suggest possible effect modification in a different direction than that found here (this would be rare); 2=there is no prior evidence mentioned or the existing evidence suggests no effect modification by this variable; 3=there is moderate prior evidence for effect modification in the direction observed: possibly from an observational study, a non-significant finding in prior RCT, or very strong biologic/pathophysiologic justification; 4=there is strong prior evidence that this variable may modify effectiveness in the direction observed; e.g., a significant interaction or a strong apparent effect in a previous, similar RCT. (Note that a risk score has strong prior evidence based both on the mathematical expectation as well as many prior studies showing that risk can modify effect, at least on the absolute scale, across many conditions and treatments). |
| D31 | | ICEMAN #3: Are any cut-points for continuous effect modifiers pre-specified? 1=cut-points are derived from the RCT data; 2=unclear; 3=cut-points suggested from prior RCT analyses; 4=variable is treated as continuous | | Using data-driven cut-points in either an effect model or risk model maximizes the probability of data over-fitting and is not at all credible. Treating the variable as continuous and showing that relationship of the variable to treatment effect is monotonic (e.g., linear or logistic) is ideal. Score as follows: 1=data-driven cut-points from exploratory analyses such as ML; 2=cutpoints appear arbitrary, rationale for these cut-points not given; 3=cut-points are pre-specified and/or come from prior RCTs; 4=variables are treated as continuous (give credit for treating the effect modifier as continuous in some analyses, even if stratified analyses are also presented). |
| D32 | | ICEMAN #4: Does a test for inter-action suggest chance is an unlikely explana-tion of the apparent effect modification?  1= interaction p-value >0.05;  2= interaction p-value between 0.05 and 0.01, OR not reported;  3=interaction p-value <0.01, but ≥0.005;  4= interaction p-value <0.005 | | Look for reports statistical tests of interaction terms in regression models, but the question of possible interaction may also be evaluated as a statistical difference between the fits of models with vs without interaction factors (e.g., likelihood ratio tests, Akaike’s Information Criterion), or by showing that the 95% confidence intervals of effect size estimtes between two or more strata of the effect modifier/covariate don’t overlap (though that would be rare).  For machine learning analyses, examine confidence intervals for effect size estimates as well as results of a variety of statistical tests of the global null hypothesis of no HTE. |
| D35 | | Credibility Score  1=very low  2=low  3=moderate  4=high | | According to ICEMAN scoring criteria, the 3 criteria above can be used to create the credibility score:  Score as “very low” (=1) if all 4 criteria are ≤ 2;  Score as “low” (=2) if at least 2 of the 4 criteria are 1 (regardless of scores for other criteria).  Score as “high) (=4) if all 4 criteria are ≥ 3;  Note: a score of “very low” or “low” may be raised to “moderate” (=3), regardless of the 3 criteria above, if the authors validated the final model presented in a fully external dataset OR in a geographically or temporal set-aside subset of original trial population.  Remaining studies could be scored as either “low” or “moderate”, depending on your assessment of the quality of the methods, extent of internal validation, the magnitude of reported heterogeneity and statistical tests for HTE or interaction (although such tests are not required for establishing the credibility of the HTE. (Note: if concerns about methods cause you to classify findings as less credible, despite high scores on ICEMAN criteria, this needs discussion among all reviewers). |
| D36 | | Credible HTE?  0=NO  1=YES | | Score as credible HTE (=1) if the final credibility score is ≥ 3 (i.e., moderate or strong); otherwise score as not credible (score = 0). |
| D38 | | Reviewer assessment of whether there is clinically important HTE? (0=NO, 1=YES, 2=UNCLEAR) | | If the report meets ICEMAN criteria for “credible HTE” then determine whether it also meets criteria for “clinically important HTE”. The PATH Statement defines this as “variation in the risk difference (i.e., the absolute treatment effect) across patient subgroups potentially sufficient to span clinically-defined decision thresholds.” Crossing a “decision threshold” should be interpreted as creating two or more patients subgroups for whom different treatment recommendations are justified. This may mean that different treatments appear superior for different subgroups OR that a treatment is clearly superior for one subgroup, but the treatment choice is not clear in another subgroup for whom preference-based decision-making may be the better choice. Clinically important HTE requires that variability be present on the risk difference (absolute) scale, even if many analyses were presented on the relative scale. “Risk magnification” (variation in absolute treatment effect with no multiplicative HTE) can be considered clinically meaningful. In making this decision, consider also whether the HTE still justifies different treatment choices in light of available information on other outcomes or patterns of adverse treatment effects. Finally, note that you are deciding whether the variation is “potentially sufficient” to span decision thresholds. This means that it does NOT require that the finding be ready for implementation, nor that the authors presented the data in a way that could be easily implemented. |
| D39 | | **For effect models only**: Do the authors report performance metrics intended for risk prediction to effect models?  0=NO  1=YES | | Some authors may treat effect models as if they are simply models that predict an individual’s risk of the outcome rather than the individual treatment effect, reporting traditional C-statistics for discrimination and/or traditional calibration curves. While this is not entirely inappropriate, it does not tell us anything about the model’s accuracy or reliability for predicting treatment effects. If such statistics are reported for the effect model, score as 1. Otherwise score as 0. NOTE: Some authors construct separate models predicting the outcome in the treatment and control arms and then calculate for each individual the ITE by subtracting predicted risk in one arm from the other. If they apply conventional C-statistics to each arm separately they are NOT applying them to an effect model. Score as 0. |
| D40 | | **For effect models only:** are performance measures specifically intended to evaluate the model’s ability to predict treatment effect or differences reported?  0=NO  1=YES | | Recently, the idea of evaluating the performance of effect models for their ability to directly predict treatment effect or differences has gained traction. Van Klaveren’s C-for-benefit is an early example that evaluates whether a treatment effect model can discriminate between persons who will benefit and those who will not from a specific treatment. A variety of approaches are presented, often in external datasets showing the model’s ability to create subgroups with predictably different average or individual treatment effects (e.g. calibration for benefit). Given the early state of this effort, we want to give credit to a variety of efforts. |

### S5. Consistency with PATH Statement criteria for risk modeling Consistency with PATH Statement criteria for risk modeling

#### Supplement Table 5. Consistency with PATH Statement criteria for risk modeling analyses (n=31)

| **Reco­­mmendation** | **Number Adherent / Number Eligible*** |
| --- | --- |
| 1. Conduct a risk model analysis if the trial has positive overall results | 25/48 (52%)^†^ |
| 2. Apply an externally developed risk model to stratify the trial population, if available | 14/31 (45%) |
| 3. If developing an internal model, avoid using control group only | 14/17 (82%)^‡^ |
| 4. Pre-specify plan for applying/developing the model | 23/31 (74%) |
| 5. Report metrics for model performance on trial population | 26/31 (84%) |
| 6. Report distribution of predicted risk for each arm of trial | 16/31 (52%) |
| 7. Report treatment effects in both relative and absolute terms across risk strata | 23/31 (77%) |
| 8. If there are important treatment-related harms (n=11), report these by risk stratum | 8/12 (67%)^§^ |
| ***** Several recommendations apply only to subset of reports, as indicated below.  ^†^ Denominator is all 48 reports from RCTs with positive overall findings.  ^‡^ Denominator excludes the 14 reports that assigned risk based on external prediction models.  ^§^ Denominator includes only those RCTs in which at least one treatment carried risks of important harms. | |

Consistency with PATH Statement criteria (Table 1) was above 60% for all but three criteria. Only half (52%) of the 48 reports with positive overall findings included a risk model. External prediction models were employed in only 14 of 31 risk model analyses, possibly because an appropriate external model was not available in the other instances; risk score distributions were presented separately by trial arm in only 16 of 31 reports. Of note, 14 of 17 reports that developed an internal risk model followed recommendations of the Statement to include both treatment arms in model development.

### S6. Consistency with PATH Statement criteria for effect modeling

#### Supplement Table 6. Consistency with PATH Statement criteria for effect modeling (41 effect models)

| **Recommendation** | **Number Adherent / Number Eligible** |
| --- | --- |
| 1. Incorporate only highly credible effect modifiers into prediction models using multiplicative interaction terms | 6/41 (15%) |
| 2. Avoid regression models that do not take into account model complexity |  |
| a. Use shrinkage methods | 34/41 (83%) |
| b. Use internal validation | 36/41 (88%) |
| c. Use external validation | 9/41 (22%) |
| 3. Avoid evaluations of treatment effect model performance that use only conventional metrics for predicting risk | 26/31* (84%) |
| 4. Report model performance in terms of ability to predict treatment effect | 14/31* (47%) |
| - Assessment of model performance is relevant only when authors claimed to have found HTE (31 of 41 effect models) | |

Most effect model reports were inconsistent with the PATH Statement criterion to include only candidate effect modifiers with prior evidence. Most did use recommended steps for reducing risks of over-fitting, including coefficient shrinkage methods and various approaches to internal validation. Only nine applied effect model findings to external datasets for validation. All but five of the 31 effect model reports that claimed to have found HTE heeded Statement advice to NOT report performance metrics designed for evaluating risk prediction when assessing their treatment effect models without also reporting performance in predicting treatment effects. In all, 14 of the 31 effect modeling reports that claimed to have identified HTE specifically assessed effect model performance for predicting individual treatment effects, i.e., by reporting discrimination statistics (e.g., C-for benefit), showing calibration graphics of observed treatment effects across the range of predicted treatment effects, or both.

### S7. Instructions for applying ICEMAN Criteria to claims of HTE

#### Supplement Table 7A. Coding Instructions for Applying Four ICEMAN Criteria to Assess Credibility of HTE^1^

1. **Did the authors test only a small number of effect modifiers or consider the number in their statistical analysis?**

**[1] Definitely no**: Explicitly exploratory analysis or large number of analyses (e.g. greater than 10) and multiplicity not considered in analysis.

[**2] Probably no or unclear**: No mention of number or 4-10 effect modifiers tested and number not considered in analysis.

**[3] Probably yes**: No protocol available but unequivocal statement of 3 or fewer effect modifiers tested.

[**4] Definitely yes**: Protocol available and 3 or fewer effect modifiers tested or number considered in analysis.

**2. Was the effect modification supported by prior evidence?**

**[1] Inconsistent with prior evidence:** Prior evidence suggested a different direction of effect modification.

**[2] Little or no support or unclear**: No prior evidence or consistent with weak or very indirect prior evidence (e.g. animal study at high risk of bias) or unclear.

**[3] Some support:** Consistent with more limited or indirect prior evidence (e.g. large observational study, non-significant effect modification in prior RCT, or different population).

**[4] Strong support:** Consistent with strong prior evidence directly applicable to the clinical scenario (e.g. significant effect modification in related RCT).

3. **If effect modifier is a continuous variable, were arbitrary cut points avoided?**

**[1] Definitely no:** Analyzed based on exploratory cut point(s) (e.g. picking cut point associated with highest interaction p-value).

**[2] Probably no or unclear:** Analyzed based on cut point(s) of unclear origin.

**[3] Probably yes:** Analysis based on pre-specified cut points, e.g. suggested by prior RCT.

**[4] Definitely yes:** Analysis based on the full continuum, e.g. assuming a linear or logarithmic relationship.

**4. Does a test for interaction suggest that chance is an unlikely explanation of the apparent effect modification?**

**[1] Chance a very likely explanation:** Interaction p-value > 0.05.

**[2] Chance a likely explanation or unclear:** Interaction p-value ≤ 0.05 and > 0.01, or no test of interaction reported and not computable.

**[3] Chance may not explain:** Interaction p-value ≤ 0.01 and > 0.005.

**[4] Chance an unlikely explanation:** Interaction p-value ≤ 0.005.

**^1^Source:** Instrument to assess the credibility of effect modification analyses (ICEMAN) in a randomized controlled trial. Available from: <https://www.iceman.help/>. Accessed on Jan 22, 2024.

#### Supplement Table 7B. Summarizing the Four Criteria Responses into an Overall Credibility Score^1^

The overall rating is a continuous visual analogue scale spanning four credibility areas. The credibility areas provide labels for credibility (the credibility areas roughly correspond to <25%, 25-50%, 50-75%, and >75% confidence that the apparent effect modification is true and not the result of chance or bias).

The overall rating should be driven by the items that decrease credibility. The following provides a sensible strategy:

1. **Very low credibility**: All responses definitely or probably reduced credibility or unclear.
2. **Low credibility (or very low):** Two or more responses definitely reduced credibility, even if all other responses satisfy credibility criteria
3. **Moderate credibility (or lower):** One response definitely reduced credibility maximum usually moderate credibility even if all other responses satisfy credibility criteria; **or:**

Two responses probably reduced credibility, maximum usually moderate credibility even if all other responses satisfy credibility criteria

4. **High credibility**: No response options definitely or probably reduced credibility 🡪 high credibility very likely

**^1^** Source**:** Instrument to assess the credibility of effect modification analyses (ICEMAN) in a randomized controlled trial. Available from: <https://www.iceman.help/>. Accessed on Jan 22, 2024.

If all 4 criteria are scored as definitely or probably credible, overall credibility of HTE is rated as “high”. If all criteria are scored as definitely or probably not credible, overall credibility is scored as “very low.” Except for these two extremes, the ICEMAN manual provides some flexibility in choosing between overall credibility scores of “low” or “moderate,” considering other aspects of findings, such as the RCT’s sample size and power for detecting interactions, whether there was consistency of effect modification across multiple outcomes, a dose-response relationship of the effect modifier with observed treatment effect, or whether findings persisted in sensitivity analyses or after adjustment for other potential effect modifiers.

**For purposes of the present analyses, reports scored as moderate or high overall credibility were classified as credible and assessed further for clinical importance.**

### S8. Results of Assessment for Overall Credibility and Clinical Importance

Most reports (51 of 65), whether of risk or effect modeling, claimed to have identified HTE. To assess credibility of claimed HTE, on either absolute or relative scales, we adapted four of the five ICEMAN criteria for RCTs (Supplement Tables 7a and 7B). Although ICEMAN criteria were originally developed for evaluating treatment effect modification by single covariates, the first four apply readily to predictive modeling with multiple covariates. These include 1) Did the authors test only a small number of interactions; 2) Was possible effect modification by each covariate supported by prior evidence; 3) If the covariate is a continuous variable, were arbitrary, data-driven cut points avoided; and 4) Does a statistical test for interaction suggest that chance is an unlikely explanation of the apparent HTE? The fifth criterion, whether the direction of interaction was hypothesized in advance, does not readily apply to multivariable predictive modeling, given that multiple and potentially complex interactions are evaluated simultaneously. According to ICEMAN guidance, no single criterion, including that of statistical testing, is treated as either sufficient or necessary for establishing overall credibility. Overall credibility scores range from 1 to 4 (very low, low, moderate, or high credibility). We used credibility scores of 3 or 4 (moderate or high credible) to assign credibility.

In Table 8 below, the 51 reports are listed first for risk modeling analyses followed by effect modeling analyses. Note that three reports (Goligher et al^84^, Harrer et al^91^, and Smit^111^) presented claimed HTE in both risk and effect models, so 54 analyses are described. Within each stratum of models, those found to be both credible and clinically important are listed first, followed by those found to be credible but not clinically important, followed by those that were judged to be not credible (and were therefore not assessed for clinical importance).

#### Supplement Table 8. Evaluation of Claimed HTE Using Four Criteria Adapted from ICEMAN* to Assess Overall Credibility of HTE and Clinical Importance

| **Ref^†^** | **Randomized Intervention(s); Patient Population; Outcomes** | **Overall RCT Findings** | **ICEMAN CRITERIA** | | | | | **ICEMAN**  **Overall Credi-bility**  **Score^‡^** | **Clinical Importance** |
| --- | --- | --- | --- | --- | --- | --- | --- | --- | --- |
|  |  |  | 1 Small # of inter-actions tested | 2. Strong prior evidence for EM | 3. No arbitrary cut points | 4. Statistical tests  support HTE | |  |  |
|  |  |  |  |  |  | Scale | Criterion Score (p-value) |  |  |
| **Risk Modeling (n=23 reports claiming HTE)** | | | | | | | | | |
| Redelmeier et al.^46,47^ | Implantable defibrillator vs medical management; patients with heart failure; all-cause mortality | Strong benefit in favor of ICD (aOR=0.69) | 4 | 4 | 3 | Rel | 4 (<0.001) | 4 | **YES**. Strong HTE on relative scale, with maximal benefit in middle of risk range and corresponding large differences in absolute risk reduction – a “sweet spot”. |
|  |  |  |  |  |  | Abs | 2 (not tested) | 3 |  |
| Chalkou et al.^48,49^ | 3 immunologic therapies (DF: Dimethyl fumarate, GA: Glatiramer acetate, N: Natalizumab) vs placebo; patients with multiple sclerosis; MS relapse | Stronger benefit for N:  (OR’s _vs placebo_:  DF: 043  GA: 0.53  N: 0.28) | 4 | 4 | 4 | Rel | 1 (>0.05) | 3 | **YES.** Suggestion of substantial HTE on both absolute and relative risk reduction. Because this is a network meta-analysis with relatively small sample sizes, 95% CI’s for both are overlapping. |
|  |  |  |  |  |  | Abs | 1 (>0.05) | 3 |  |
| Kumar et al^54^ | Apixaban vs placebo; ambulatory patients with cancer; venous thrombo-embolism (VTE) | Strong benefit in favor of apixaban (aHR=0.49) | 4 | 4 | 3 | Rel | 1 (>0.05) | 2 | **YES**. Large subgroup differences in both relative and absolute risk reduction. Small trial size (n=466), very small high-risk sub-group (n=155) resulted in neither difference reaching statistical significance. |
|  |  |  |  |  |  | Abs | 1 (>0.05) | 3 |  |
| Rysavy et al^59^ | Vitamin A vs sham injection; infants with extreme prematurity; bronchopulmonary dysplasia | Weak benefit in favor of vitamin A (aRR=0.89) | 4 | 4 | 3 | Rel | 2 (<0.05) | 3 | **YES.** Quintile specific analyses suggest substantial HTE, with benefit confined to lower-risk infants; p-value for multiplicative interaction <0.05, although 95% CI’s appear to overlap for both OR and RD |
|  |  |  |  |  |  | Abs | 1 (>0.05) | 3 |  |
| Bress et al.^60^ | Intensive vs standard systolic blood pressure (SBP) control; patients with systolic hypertension and increased cardiovascular disease (CVD) risk; CVD events, all-cause mortality | Strong benefits in favor of intensive control (aHR=0.75 and 0.73, respectively) | 4 | 4 | 4 | Rel | 2 (tested but not reported) | 2 | **YES.** Risk magnification, with differences in ARR large enough to be important; because an interaction term on relative scale is included in the model but results are not shown, a possible difference in relative treatment effects across baseline risk cannot be excluded. |
|  |  |  |  |  |  | Abs | 1 (>0.05) | 3 |  |
| Kent et al^61^ | Percutaneous PFO-closure vs medical therapy; patients with a patent foramen ovale (PFO) - associated stroke; recurrent stroke | Strong benefit in favor of PFO-closure (aHR=0.41) | 4 | 4 | 3 | Rel | 4 (0.003) | 4 | **YES**. Strong multiplicative interaction (qualitative) with parallel, large subgroup differences in absolute risk reduction. |
|  |  |  |  |  |  | Abs | 1 (>0.05) | 3 |  |
| Gencer et al.^68^ | Lower (LDER) vs higher (HDER) dose edoxaban vs warfarin (WARF); patients with atrial fibrillation;  composite of stroke/  systemic embolism, major bleed, all-cause mortality | Weak benefit for both HDER and LDER vs. WARF (HR=0.83 for LDER 0.89 for HDER | 4 | 4 | 3 | Rel | 1 (0.43) | 1 | **YES**. Clear risk magnification with statistical tests that confirm the visual presentation; no suggestion of differences in relative treatment effects across baseline risk. |
|  |  |  |  |  |  | Abs | 4 (<0.001) | 4 |  |
| Taylor et al.^69^ | Nurse-navigator led sepsis transition and recovery intervention vs usual care; hospitalized patients with sepsis; 30-day mortality and readmission | Weak benefit in favor of the inter-vention (aOR=0.80) | 4 | 4 | 3 | Rel | 2 (<0.05) | 3 | **YES**. 95% CI’s for quartile-specific treatment effects are non-overlapping on both rel and abs scales, with maximal benefit in the middle of the risk range – a “sweet spot.” |
|  |  |  |  |  |  | Abs | 1 (>0.05) | 3 |  |
| Trinks-Roerdink et al^79^ | Integrated atrial fibrillation care vs. usual care; patients with atrial fibrillation; all-cause mortality | Strong benefit of integrated care (aHR=0.55) | 4 | 4 | 3 | Rel | 1 (>0.05) | 1 | **YES.** Appearance of clear risk magnification. Increasing absolute risk reduction with increasing risk, although 95% CIs of extreme quartiles overlap slightly; no interaction on relative scale. |
|  |  |  |  |  |  | Abs | 2 (not tested) | 3 |  |
| Goligher et al ^81^ | Therapeutic-dose heparin vs usual pharmacologic thromboprophylaxis; hospitalized patients with Covid-19 infection; organ-support free days, hospital survival | No benefit in overall population (OR for benefit 1.05) | 4 | 4 | 3 | Rel | 2 (not reported) | 3 | **YES.** Apparent qualitative interaction on both rel and abs scale with a gradient (decreasing benefit with increasing risk). However, ARRs are small everywhere (see eTable 5 in supplement to publication); |
|  |  |  |  |  |  | Abs | 2 (0.05) | 3 |  |
| Paules et al^96^ | Baricitinab + remdesivir vs. remdesivir alone; patients hospitalized with Covid 19; 28-day mortality; progression to mechanical ventilation or death; recovery by 28-days. | Strong benefit of adding Baricitinib for all 3 outcomes  (OR for 28-day mortality: 0.65) | 4 | 4 | 3 | Rel | 2 (<0.05) | 3 | **YES.** Clear increases in absolute benefit for all 3 outcomes with increasing risk. Probable multiplicative interaction for recovery rate, with highest risk quartile having a greater relative effect; risk magnification for the other 2 outcomes |
|  |  |  |  |  |  | Abs | 2 (p<0.05) | 4 |  |
| Vickers et al^97^ | Radical prostatectomy vs conservative care; patients with localized prostate cancer; 15-year prostate cancer mortality | Strong Benefit (aHR= 0.60; 95% CI 0.37- 0.99 | 4 | 4 | 3 | Rel | 1 (p>0.05) | 1 | **YES.** Clear, strong risk magnification with no suggestion of differences in relative effects by risk quartile |
|  |  |  |  |  |  | Abs | 2 (not tested) | 4 |  |
| De Winkel et al^106^ | Endovascular coiling vs intracranial clip; patients with acute subarachnoid hemorrhage; functional status at 2 mos; re-treatment or re-bleed by 10 yrs. | Results strongly favor coiling for short-term functional status, but clipping for long-term re-bleeding/re-treatment | 4 | 3 | 4 | Rel | 1 (p>0.05) | 1 | **YES.** Started as effect models, but found no relative HTE for either outcome; converted to risk models for each outcome and identified a small subgroup (6%) in which long-term benefit of clipping was large, but short-term benefit of coiling was small. |
|  |  |  |  |  |  | Abs | 2 (not tested) | 4 |  |
| Xu et al^94^ | low-dose aspirin vs placebo; healthy people aged 70 and above; composite of death, dementia or disability | Weak benefit of aspirin (HR=0.85) | 4 | 4 | 3 | Rel | 1 (p> 0.05 | 1 | **YES.** Risk stratification demonstrated strong benefit on absolute scale only in the highest risk quintile. Little benefit elsewhere. |
|  |  |  |  |  |  | Abs | 4 (p=0.03) | 3 |  |
| Smit et al^111^ | Corticosteroids vs placebo; patients with community-acquired pneumonia; 30-day mortality | Strong benefit of corticosteroids (OR 0.72; 95% CI .56-0.94) | 4 | 4 | 4 | Rel | 1 (p=0.54) | 1 | **YES.** Strong HTE on absolute scale; possibly also on relative scale. Despite overall benefit in prior meta-analyses, risk modeling shows that 60% of low-risk patients would not benefit. |
|  |  |  |  |  |  | Abs | 2 (p<0.05) | 3 |  |
| Van Kruijsdijk et al.^72^ | Hemodiafiltration vs hemodialysis ; chronic renal failure; all-cause mortality, median survival | Strong benefit for all-cause mortality (HR= 0.78) | 4 | 4 | 3 | Rel | 1 (P=0.63) | 1 | **NO.** clear risk magnification demonstrated, but because authors stratified on probability of long-term survival rather than mortality, findings are subject to multiple competing risks and are not clinically useful. |
|  |  |  |  |  |  | Abs | 2 (not reported) | 3 |  |
| Mell et al^76^ | Androgen deprivation vs control; prostate cancer (PC); competing events: PC-related event (distant metastasis or death) vs non-PC death, overall mortality | Strong benefit for PC-related event (HR=0.61); moderate benefit for all-cause mortality (HR=0.82) | 4 | 4 | 4 | Rel | 2 (p<0.05) | 3 | **NO.** Significant multiplicative HTE and apparent absolute increases in treatment effect with higher baseline risk. However the risk score developed in the paper did not appear to add to or do as well as current risk stratification strategies for identifying HTE |
|  |  |  |  |  |  | Abs | 2 (not reported) | 3 |  |
| Li^92^ | Early introduction of peanuts vs peanut avoidance; infants at high risk for peanut allergy; peanut allergy at 5 yrs | Very strong benefit of early introduction (RR=0.14, p<0.001) | 4 | 4 | 4 | Rel | 1 (p>0.05) | 1 | **NO.** Risk magnification is demonstrated, but benefit is substantial even in lowest risk stratum. No suggestion of a decision threshold. |
|  |  |  |  |  |  | Abs | 4 (p<0.005) | 4 |  |
| Burger^112^ | Icosapent ethyl vs placebo; patients with cardiovascular disease or high risk; major adverse cardiovascular events (3-point MACE) | Strong benefit:  ((HR, 0.75; 95% CI 0.68-0.83; | 4 | 4 | 4 | Rel | 1 (p>0.05) | 1 | **NO.** Modest risk magnification demonstrated for 3-point MACE, but sizeable ARR even in lowest-risk quartile. No suggestion of a decision threshold. |
|  |  |  |  |  |  | Abs | 3 (not tested) | 3 |  |
| Charu et al^85^ | Intensive vs. standard glycemic control; diabetes mellitus; severe microalbuminuria or kidney failure; all-cause mortality | Strong benefit for severe microalbuminuria (HR = 0.75, 95% CI 0.65-0.86) | 4 | 4 | 3 | Rel | 2 (not tested) | 2 | **NO**; although risk magnification is seen using one risk score, effects on microvascular outcomes conflict with results for all-cause mortality; effects limited to reduction in new onset albuminuria; |
|  |  |  |  |  |  | Abs | 2 (not tested) | 3 |  |
| Brade et al^62^ | Various performance feedback interventions; newly enrolled undergraduate university students; academic performance outcomes (credits obtained; grade point average; graduation rates) | 3 RCTs reported. Most overall effects null. One RCT finds weak benefit (aRR=1.10) on graduation rates. | 2 | 2 | 3 | Rel | 2 (p<0.05) | 2 | **NO**; multiple interventions and outcomes tested for HTE; risk scores not well defined. Multiple tests of HTE. Findings difficult to interpret or apply. |
|  |  |  |  |  |  | Abs | 2 (not tested) | 1 |  |
| Ghazi et al^83^ | Intensive vs standard systolic blood pressure (SBP) control; patients with high SBP and increased cardiovascular disease (CVD) risk; probable dementia or mild amnestic cognitive impairment. | Weak benefit (crude risk ratio calculated from Table 2: 0.91, p<0.05) | 4 | 4 | 4 | Rel | 2 (not reported) | 1 | **NO**; results not interpretable due to incomplete reporting; marginal overall benefit; unclear whether there is risk magnification or HTE on the relative scale with possible reduced ARR at highest levels of risk. |
|  |  |  |  |  |  | Abs | 2 (not tested) | 2 |  |
|  |  |  |  |  |  | Abs | 2 (not tested) | 3 |  |
| Harrer^91^ | Digital (i.e., online) stress reduction intervention vs waitlist; patients with clinically relevant symptoms of depression; CES-D score at 6 mos. | Weak benefit: standard effect size -0.56; 95% CI 0.37-75 | 2 | 2 | 3 | Rel | 2 (<0.05) | 2 | **NO.** A strong interaction of treatment with a risk score is claimed, with stronger effects in the higher risk half of population, but risk score is poorly described; and treatment effect differences across the two risk strata are not shown. |
|  |  |  |  |  |  | Abs | 2 (not reported) | 1 |  |
| **Effect Modeling (n=31 reports claiming HTE)** | | | | | | | | | |
| Park et al^51^ | Convalescent plasma (CCP) vs control; hospitalized patients with Covid-19 without mechanical ventilation; ordinal COVID-19 clinical status scale | Null (no  overall association between CCP and patient outcomes) | 2 | 2 | 4 | Rel | 3(<0.01) | 3 | **YES**. Fails two criteria, but large relative and absolute differences between extreme tertiles of predicted benefit demonstrated in multiple external RCT database validations |
|  |  |  |  |  |  | Abs | 2 (not reported) | 3 |  |
| Takahashi et al^52^ | Coronary artery bypass (CABG) vs. percutaneous coronary intervention, patients with new 3-vessel or left main coronary artery disease, 5-year major adverse cardiac events (MACE) and 10-yr all cause death. | Weak benefit favoring CABG (HR for 10-yr survival 1.19, 95% CI 0.99-1.43) | 4 | 4 | 3 | Rel | 2 (<0.05) | 3 | **YES**. All criteria met; and model predictions for individual treatment effects were also externally validated and reproduced, including large, nearly significant subgroup differences in ARRs (see Fig 4 in publication). |
|  |  |  |  |  |  | Abs | 1 (>0.05) | 3 |  |
| Dennis et al.^70^ | Initiation of SGLT-2 inhibitors vs DPP-4 inhibitor as add-on therapy, patients with Type 2 diabetes  Hb A1c at 6 mos post-initiation | Weak overall benefit in favor of SGLT-2 inhibitors; Mean A1c difference: 1.2mmol.mol | 1 | 2 | 4 | Abs | 2 (<0.05) | 3 | **YES**. Initial effect model fails ICEMAN criteria 1 and 2; but multiple external validations reproduced model treatment effect predictions with p-values below 0.05 for subgroup differences, identifying groups that did better with SGLT-2 inhibitors, a smaller group that did better if a DPP-4 agent was initiated. |
| Seitz et al^80^ | Bougie vs Stylet for intubation; patients with respiratory distress; success on first attempt at intubation | Null (non-significant 6.8 percent point difference in favor of stylet) | 1 | 2 | 1 | Abs | 2 (<0.05) | 3 | **YES**. Fails first three criteria, but model developed in one cohort; evaluated in independent RCT cohort; p-values are from external validation cohort. |
| Venkatasubramaniam et al^92^ | SGLT-2 vs DPP-4i’s as add-on therapy; patients with non-insulin-using T2 diabetes; Hb A1c reduction at 6 mos | Weak benefit favoring SGLT-2i’s | 1 | 2 | 3 | Abs | 4 (p<0.005) | 3 | **YES.** Exploratory approach would not meet ICEMAN criteria, but model was externally validated, identifying a group (11%) who do better on DPP=4is |
| Buell et al^93^ | Lower vs higher 02 saturation (Spo_2_) targets; Patients requiring mechanical ventilation; 28-day mortality | No significant difference between the two treatments | 1 | 2 | 1 | Abs | 2(p<0.05) | 3 | **YES.** Exploratory approach does not meet ICEMAN criteria, but model was externally validated, with findings of significant HTE, and identifying 33% of patients benefitting from lower Spo_2_ and 33% benefitting from higher Spo_2_ targets |
| Arnold et al^99^ | Coronary revasculariza-tion vs medical therapy; patients with chronic coronary artery disease; Seattle Angina Questionnaire summary score (SAQ-SS) at 12 mos | Weak benefit: 4.2 point difference in SAQ-SS at 12 mos (95% CI 3.3-5.1) | 3 | 3 | 3 | Rel | 2 (p<0.03) | 3 | **YES.** An effect model with a primary focus on a single interaction. Meets ICEMAN criteria and identifies a subgroup who clearly benefits and another for whom shared decision-making is most appropriate |
|  |  |  |  |  |  | Abs | 2 (<0.05) | 3 |  |
| Ninomiya et al^108^ | Coronary artery bypass grafting (CABG) vs. percutaneous coronary intervention (PCI); patients with 3-vessel or left main coronary artery disease; 5-yr all-cause mortality | Weak benefit favoring CABG (HR for 10-yr survival 1.19, 95% CI 0.99-1.43) | 4 | 4 | 4 | Rel | 3 (<0.01) | 4 | **YES.** All ICEMAN criteria met, and model findings validated externally. Identifies a subgroup of 50% who clearly benefit from CABG; 25% who do better with PCI. |
|  |  |  |  |  |  | Abs | 2 (<0.05) | 4 |  |
| Smit et al^111^ | Corticosteroids vs placebo; patients with community-acquired pneumonia; 30-day mortality | Strong benefit (OR 0·72; 95% CI 0·56-0·94) | 1 | 2 | 4 | Rel | 2 (0.03) | 3 | **YES.** Initial effect model fails ICEMAN criteria; but an external validation replicated the ability of the model to identify a large subgroup with low CRP levels and no apparent benefit. |
|  |  |  |  |  |  | Abs | 2(<0.05) | 3 |  |
| Dennis et al^55^ | Metformin (MET) vs sulfonylurea (SU) vs thiazolidinedione (TZD) as initial therapy; type 2 diabetes mellitus; failure of glycemic control during follow-up. | Strong benefits of TZD vs SU (RR=0.44) and vs MET (RR=0.71) (ADOPT trial) | 2 | 2 | 4 | Abs | 2(not reported) | 3 | **NO**. Although predictive model appears to perform better than treatment choice based on clusters, details are limited and authors conclude that findings are not yet ready for clinical use. |
| Nguyen et al (2020)^53^ | Aspirin therapy; patients with acute ischemic stroke; death or dependency at 6 months | Weak benefit: ARR: -1.3% (95% CI: -2.5% - 0.1%) | 1 | 2 | 4 | Rel | 2 (not tested) | 1 | **NO**. largely a methodologic study of an exploratory approach involving 23 potential effect modifiers. No information provided on key effect modifiers among them. External validation given in supplement does not suggest strong predictive ability for individual treatment effect. |
|  |  |  |  |  |  | Abs | 2 (p<0.05) | 2 |  |
| Rudolph et al (2021)^56^ | Treatment rule for choosing XR-Naltrexone vs buprenorphine; opioid use disorder; substance use relapse | Probable weak overall benefit of buprenorphine vs XR-naltrexone (RR=0.87) | 1 | 2 | 1 | Abs | 2 (not reported) | 1 | **NO.** HTE not credible. Exploratory findings, multiple potential interactions tested. No external validation. Absolute risk differences are not presented by subgroups defined by the treatment rule. Model does not appear to perform better than a rule based on a single variable: homelessness. |
| Fazzari et al^57^ | Individual vs group acupuncture; chronic pain; 30% reduction in pain index at 12 weeks. | Non-inferiority trial failed to establish non-inferiority of group vs individual acupuncture | 2 | 2 | 1 | Abs | 2 (p<0.05) | 1 | **NO**. HTE not credible. Exploratory findings, multiple potential interactions tested. No external validation. Additional doubts related to apparent lack of efficacy for either treatment in largest subgroup. |
| Reinhardt et al^63^ | Dabigatran (110 or 150mg) vs warfarin; atrial fibrillation; stroke or systemic embolism, major bleeding | Strong effect favoring dabigatran 150mg vs warfarin for primary outcome: RR= 0.66; 95% CI, 0.53 to 0.82 | 1 | 2 | 1 | Rel | 2 (not reported) | 1 | **NO**. HTE not credible. Exploratory findings, multiple potential interactions tested. HTE not credible. No external validation. Details of absolute risk reductions not given for subgroups identified by model. |
|  |  |  |  |  |  | Abs | 2 (not reported) | 1 |  |
| Kessler et al^64^ | Treatment rule for continuing sertraline vs switching to mirtazapine vs combining both as 2^nd^ line treatment; new onset major depression with initial sertraline failure; remission at 9 wks | Strong effect favoring combination, with switching second best; point estimates for treatment effects not given | 1 | 2 | 1 | Abs | 2 (not reported) | 1 | **NO**. HTE not credible. Exploratory findings, multiple potential interactions tested. HTE not credible. No external validation. Absolute risk differences not presented for subgroups defined by the treatment rule. |
| Edward et al^66^ | Intensive glycemic control; type 2 diabetes; major adverse cardiovascular events. | No significant overall treatment effect | 1 | 2 | 1 | Abs | 2 (<0.05) | 1 | **NO.** HTE not credible. Exploratory findings, multiple (>40) potential interactions tested. No external validation and lack of consistency between the two RCTs in this IPDMA |
| Chen et al^73^ | Lower vs higher tidal volume during ventilation; acute respiratory distress syndrome; days to returning home in survivors | Strong effect: -24 days reduction in time to return home, (95% CI : 17-35) in lower tidal volume arm) | 1 | 2 | 1 | Abs | 1 (>0.05) | 1 | **NO**. HTE not credible. Largely a methodologic study of analyzing treatment effects in cohorts with large proportions of truncated follow-up. The overall variation in estimated treatment effect appears small, unidirectional, without statistical significance across the range. |
| Wolf et al^74^ | Immediate vs gradual nicotine reduction; current smokers; change in biomarkers; average number of cigarettes smoked per day. | Strong effect on biomarker outcomes favoring immediate vs gradual reduction; p<0.01 | 1 | 2 | 1 | Abs | 2 (<0.05) | 1 | **NO.** HTE not credible. Largely a methodologic study. Exploratory findings, multiple (40) potential interactions tested. No external validation. Subgroup differences in absolute treatment effect quite small. |
| Nguyen et al (2022)^76^ | Pravastatin vs placebo; elderly high-risk patients risk for vascular disease; major adverse cardiovascular events | Weak benefit: HR 0.85, 95% CI 0.74-0.97 | 1 | 2 | 1 | Rel | 2 (<0.05) | 1 | **NO.** HTE not credible. Exploratory findings, multiple (28) potential interactions tested. No external validation |
| Rudolph et al (2023)^78^ | Buprenorphine vs methadone vs naltrexone; opioid use disorder; return to regular opioid use. | Probable weak benefit of Buprenorphine vs either methadone or naltrexone : p<0.05 | 1 | 3 | 1 | Abs | 2 (not tested) | 2 | **NO.** HTE not credible. Exploratory findings, multiple (>10) potential interactions tested; no external validation. Effect size differences between subgroups not reported. The treatment rule does not appear to improve on buprenorphine for all. Major differences in populations and treatments of the 3 RCTs included in this IPDMA. |
| Colloca et al^83^ | Tanezumab vs placebo; moderate to severe osteoarthritis; pain indices at 16 weeks. | Tanezumab superior to placebo for pain outcomes in pre-approval trials (p<0.01), but concerns raised regarding adverse events | 1 | 2 | 1 | Abs | 2 (not tested) | 1 | **NO.** HTE not credible. Exploratory findings, multiple (22) potential interactions tested. No external validation. No clinical utility as tanezumab has not been approved for use in osteoarthritis due to potential adverse effects. |
| Goligher et al^84^ | Therapeutic-dose heparin vs usual pharmacologic thromboprophylaxis; hospitalized patients with Covid-19 infection; organ-support free days, hospital survival | No benefit in overall population (OR for benefit 1.05) | 3 | 2 | 3 | Abs | 2 (p<0.05) | 2 | **NO.** HTE not credible for effect model. Did not appear to add information to that provided by the risk model in the same dataset. Subgroup differences in observed absolute risk reduction across deciles of predicted benefit were small. |
| Inoue et al^85^ | Intensive SBP control; adults with increased risk for cardiovascular disease; major adverse cardiovascular events | Strong benefit of intensive control in non-diabetic patients: HR: 0.73; non-significant benefit in diabetic patients: HR: 0.88 | 1 | 2 | 1 | Abs | 2 (p<0.05) | 1 | **NO.** HTE not credible. Exploratory findings, multiple (>10) potential interactions tested. No external validation. Findings conflict with other studies from same RCTs that did not find HTE in effect models. |
| Zarski et al^89^ | Internet-based intervention vs wait-list control; genito-pelvic pain/penetration-disorder; multi-dimensional symptom score at 12 weeks post-intervention | Weak improvement in female sexual functioning with intervention (p<0.05) | 1 | 2 | 1 | Abs | 3 (p<0.01) | 2 | **NO.** HTE not credible. Exploratory findings, multiple (39) potential interactions tested. No external validation. |
| Harrer et al^90^ | Internet-based cognitive behavioral therapy intervention vs treatment as usual; patients with depression and chronic back pain; depression severity at 9 weeks | Strong benefit favoring the intervention: (p<0.01 ) | 1 | 2 | 1 | Abs | 2 (p<0.05) | 2 | **NO.** HTE not credible. Exploratory findings, multiple (20) potential interactions tested. A small pilot external validation is mentioned but details not provided. Model does not convincingly identify subgroups that should not be treated. |
| Harrer et al^91^ | Digital (i.e., online) stress reduction intervention vs waitlist; patients with clinically relevant symptoms of depression; CES-D score at 6 mos. | Weak benefit of the intervention: Standard effect size -0.56; 95% CI 0.37 -0.75 | 1 | 2 | 4 | Abs | 2 (p<0.05) | 2 | **NO.** HTE not credible. Multiple potential interactions tested. No external validation presented. |
| Zhou et al^98^ | CABG vs medical therapy alone; patients with ischemic cardiomyopathy; 10-year all-cause mortality | Weak benefit: HR: 0.84; 95% CI 0.73 - 0.97 | 1 | 2 | 1 | Rel | 2 (not tested) | 2 | **NO.** HTE not credible. Multiple potential interactions tested. HTE not clearly established. No external validation. |
|  |  |  |  |  |  | Abs | 2 (not tested) | 2 |  |
| Desai et al^101^ | Spironolactone vs placebo; patients with heart failure & preserved EF; restricted mean survival time at 3.3 years; composite of CVD death, aborted cardiac arrest or hospitalization for Hf | Weak benefit: HR: 0.89; 95% CI 0.77 - 1.04 | 1 | 1 | 1 | Abs | 1 (p>0.05 | 1 | **NO.** HTE not credible. Multiple interactions tested. No sizeable or significant subgroup differences in treatment effects, no significant treatment-predictor interactions or measures of overall HTE. |
| Hamaya et al^103^ | 2 x 2 trial, comparing average vs high-protein, and low vs high-fat diets for wt reduction;  overweight/obese adults; 2-yr wt change | Null overall findings for both comparisons | 1 | 1 | 1 | Abs | 1 (p>0.05) | 1 | **NO.** Highly exploratory, methodologic study presenting various machine-learning approaches. No overall effects of either diet, and no HTE claimed for high protein. Possible HTE found for high-fat diet, but not credible. No external validation. |
| Afshar et al^104^ | Extended release - naltrexone vs. buprenorphine/naloxone; patients with opioid use disorder; abstinence at 6 mos | Weak benefit of buprenorphine/naloxone  (RR=0.88) | 1 | 2 | 1 | Rel | 2 (<0.05) | 1 | **NO.** HTE not credible. Multiple candidate effect modifiers examined. No external validation. |
|  |  |  |  |  |  | Abs | 1 (>0.05) | 1 |  |
| Huang et al ^106^ | chiglitazar vs sitagliptin; patients with Type 2 diabetes; Hb A1c reduction at 24 weeks | Null: no differences in outcomes | 1 | 1 | 1 | Abs | 2 (not reported) | 1 | **NO.** HTE not credible. Highly exploratory analyses; no validation. |
| *** Source:** Instrument to assess the credibility of effect modification analyses (ICEMAN) in a randomized controlled trial. Available from: <https://www.iceman.help/>. Accessed on Jan 22, 2024.  **^†^** Reference numbers refer to bibliography in the main paper.  **^‡^** Overall credibility scores of 3 or 4 were counted as “credible HTE”. | | | | | | | | | |
